# Continuous antiretroviral therapy induces progressive senescence-like reprogramming of alveolar macrophages

**DOI:** 10.1101/2025.04.02.25325039

**Authors:** Vinicius M. Fava, Monica Dallmann-Sauer, Marianna Orlova, Wilian Correa-Macedo, Ron Olivenstein, Cecilia Theresa Costiniuk, Jean-Pierre Routy, Luis B. Barreiro, Erwin Schurr

## Abstract

**Background:** Advances in antiretroviral therapy (ART) have substantially improved the lives of people with HIV (PWH) and reduced HIV acquisition through pre-exposure prophylaxis (PrEP). However, the long-term effect of ART on the physiological state of cells remains poorly understood. Despite the success of ART in preventing the progression of HIV infection to AIDS, aging PWH are suffering from a disproportional burden of non-AIDS comorbidities, including lung diseases.

**Methods:** Given the central function of alveolar macrophages (AM) in pulmonary immunity, we evaluated the impact of nucleoside reverse transcriptase inhibitors (NRTI) based ART on AM of PWH and people on PrEP. We employed a retrospective cross-sectional design and evaluated the effects of continuous exposure to ART in the epigenetic and transcript of AM at bulk and single-nucleus multiomics levels.

**Results:** We showed that continuous NRTI-based ART induces a progressive senescence-like pro-inflammatory state in AM, characterized by increased constitutive epigenetic and transcriptomic priming of genes involved in cell cycle arrest (e.g., *CDKN1A*/p21), components of the senescence-associated secretory phenotype (e.g., *IL1A*, *IL6R,* and *CXCL8*), as well as transcription factors subunits of the AP-1 family (e.g., *FOSL1, JUN*). At the AM single nucleus level we discovered a coordinated gene regulatory network linking key pro-inflammatory transcription factors to the transcriptomic alterations induced by ART. The senescence ART-linked epigenetic and transcriptomic changes were strongly dependent on the duration of ART and irrespective of HIV infection. A secondary time independent ART effect was observed for interferon signaling genes. This effect was more pronounced in people on PrEP and impaired the AM response to *ex vivo* challenge with SARS-CoV-2.

**Conclusion:** Combined, our data indicated that continuous NRTI-based ART promoted a dysregulated physiological state in AM, potentially contributing to age-related pulmonary comorbidities in PWH, such as COPD, pulmonary fibrosis, and asthma. The results of our study advocate for optimized or adjuvant therapies to mitigate potential long-term adverse effects of ART for people on PrEP and PWH.

## Background

Advances in antiretroviral therapy (ART) have improved the quality of life for people with HIV (PWH). In contrast to the initial regimens, current ART is more effective in the suppression of HIV and has lower toxicity and adverse effects (1). In addition to treatment and health care improvements, mitigation of opportunistic infections, and management of HIV comorbidities have greatly increased life expectancy for PWH (2). Consequently, HIV infection is nowadays considered a chronic manageable condition. The increased lifespan of PWH coincided with a surge of age-related idiopathic diseases despite well-controlled HIV levels (3, 4), and the aging population of PWH now suffers a disproportional burden of cardiovascular, neurological, and pulmonary diseases (5–7). The reasons for the increased incidence of non-AIDS diseases in PWH remain poorly understood. HIV persistence, low levels of inflammation and uncharacterized adverse effects of ART are among the potential contributors leading to idiopathic diseases in PWH (8–12).

With respect to pulmonary involvement, PWH receiving ART exhibit increased susceptibility to an array of pulmonary pathologies, including chronic obstructive pulmonary disease (COPD) (13), mortality and severity due to COVID-19 (14, 15), tuberculosis (TB) (16, 17), pneumonia (18, 19), pulmonary fibrosis (20), and lung cancers (21, 22). Central to the pathogenesis of these pulmonary comorbidities are alveolar macrophages (AM) which are long-lived and self-renewing sentinel cells within the lung microenvironment. We previously showed that AM isolated from PWH displayed an attenuated epigenetic and transcriptomic response to *Mycobacterium tuberculosis* (*Mtb*), the etiological agent of TB (17). Surprisingly, HIV-negative individuals receiving ART as pre-exposure prophylaxis (PrEP) similarly exhibited attenuated AM responses to *Mtb*, implicating a direct pharmacologic effect of ART independent of HIV status. Accordingly, we sought to delineate the impact of ART on the epigenetic and transcriptomic landscape of AMs in both PWH and persons receiving PrEP. We found that exposure to ART significantly altered the epigenetic and transcriptomic state of AM, imprinting a progressive, duration-dependent senescence-like phenotype and a disruption of constitutive interferon signaling. To further characterize the effects of ART on AM heterogeneity, we employed a single-nucleus multiomics (snMulti) approach to assess AM responses following SARS-CoV-2 challenge. These experiments revealed that ART modulates the composition of AM subpopulations and substantially diminishes AM *ex vivo* reactivity toward SARS-CoV-2.

## Methods

### Subjects and study design

To study the impact of ART on the AM physiological state, we used both bulk and snMulti approaches (**Fig. 1**). Subjects included in the bulk section of this study were enrolled as part of our study on AM responses to *Mtb*. Hence, details of quality control regarding Bronchoalveolar Lavages (BAL) practice, AM processing and preparation were as described previously (17). We excluded subjects with previous pulmonary infections, abnormalities in baseline lung functions, pulmonary conditions, chronic cardiovascular or metabolic diseases and illegal substances abuse. In addition, we applied the following technical exclusion criteria for the bulk approach: samples with fraction of reads in peaks < 10% and libraries with less than 20 million reads. Of the 42 subjects that passed quality control, 14 were healthy HIV-negative without ART (HC); 10 were HIV-negative at a high exposure setting for HIV under PrEP; and 18 were PWH under ART (**Table 1**). Participants had uninterrupted use of ART ranging from two months to 5.4 years for PrEP and from seven months to 23.1 years for PWH at the time of BAL cell collection. None of the participants had developed resistance to ART and all regimens included at least one NRTI. Due to the limited yield of AM from BAL some bulk assays could not be tested in all subjects (**Table 1**). For the snMulti approach we enrolled 3 PrEP and 3 PHW participants on long term ART, and 3 HC for BAL recollection (**Table 1**). However, one HC sample failed library preparation and was excluded.

**Fig. 1.**
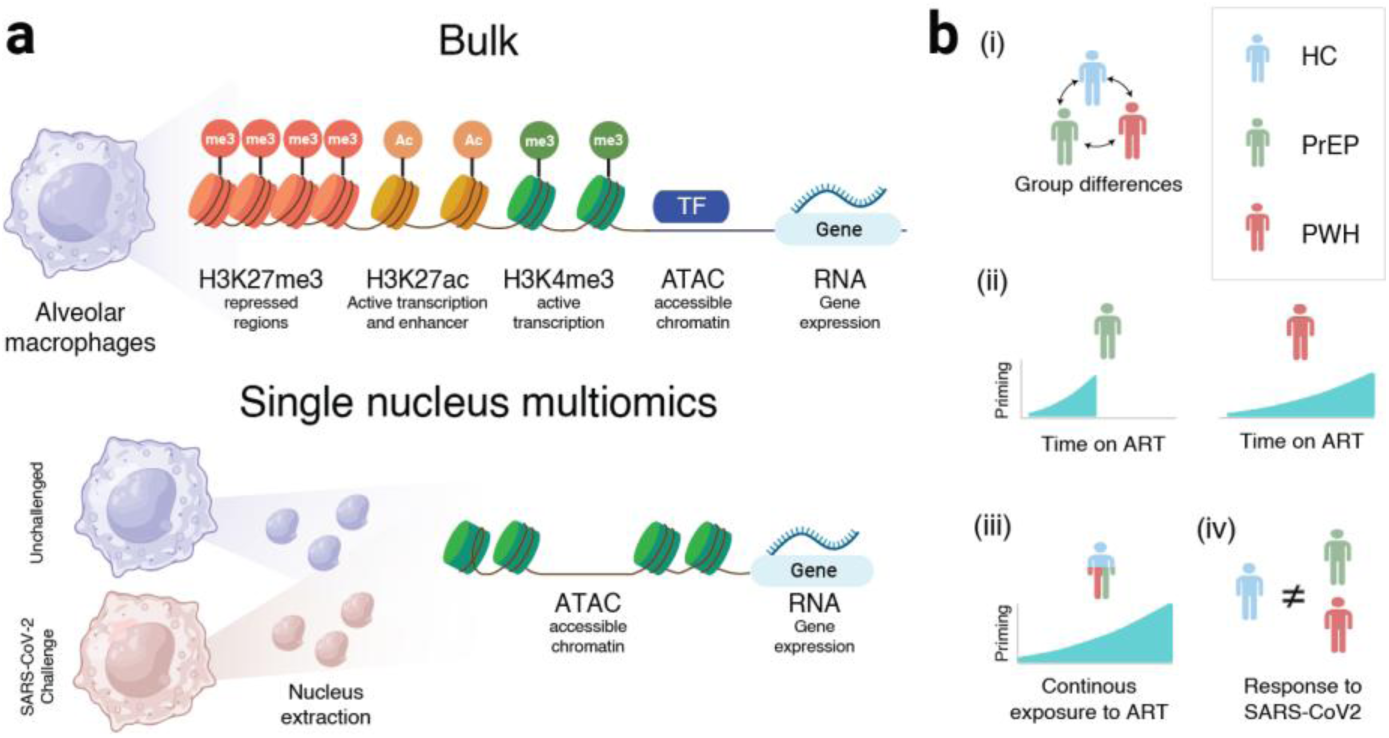
Study design overview. (a) Assays used to evaluate the physiological state of alveolar macrophages at bulk and single nucleus level. (**b**) Models tested to assess the biological mechanisms affected by ART in alveolar macrophages: (i) Differences between groups independent of duration of time on ART; (ii) The effect of time on ART for PrEP and PWH participants; (iii) Continuous exposure to ART while accounting for group status, (iv) Group specific difference in response to SARS-CoV2 challenge. (Created with BioRender).

**Table 1.** Characteristics of enrolled subjects.

| Sample | Group | Sex | Age | Smoker<sup>a</sup> | ART regimen | Molecules | Duration of HIV | Time on ART<sup>b</sup> | RNA | ATAC | H3K27ac | H3K27me3 | H3K4me3 | snMulti |
| --- | --- | --- | --- | --- | --- | --- | --- | --- | --- | --- | --- | --- | --- | --- |
| AMC2 | HC | M | 56 – 60 | YES | . | . | . | 0 | X | X | . | . | . | . |
| AMC7 | HC | M | 46 – 50 | NO | . | . | . | 0 | X | X | . | . | . | . |
| AMC8 | HC | M | 21 – 25 | NO | . | . | . | 0 | X | . | . | . | . | . |
| AMC9 | HC | M | 56 – 60 | YES | . | . | . | 0 | X | X | X | X | X | . |
| AMC10 | HC | M | 26 – 30 | NO | . | . | . | 0 | X | X | X | . | X | . |
| AMC11 | HC | M | 26 – 30 | NO | . | . | . | 0 | X | X | X | . | . | . |
| AMC12 | HC | M | 35 – 40 | NO | . | . | . | 0 | X | X | X | . | X | . |
| AMC12.2<sup>c</sup> | HC | M | 41 – 45 | NO | . | . | . | 0 | . | . | . | . | . | X |
| AMC13 | HC | F | 35 – 40 | NO | . | . | . | 0 | X | X | X | X | X | . |
| AMC14 | HC | F | 35 – 40 | YES | . | . | . | 0 | X | X | . | X | . | . |
| AMC15 | HC | F | 41 – 45 | NO | . | . | . | 0 | X | X | . | . | . | . |
| AMC16 | HC | F | 35 – 40 | NO | . | . | . | 0 | X | X | . | . | . | . |
| AMC18 | HC | M | 26 – 30 | NO | . | . | . | 0 | X | X | . | . | . | . |
| AMC19 | HC | M | 51 – 55 | NO | . | . | . | 0 | X | X | . | . | . | . |
| AMC22 | HC | F | 41 – 45 | YES | . | . | . | 0 | X | X | X | X | X | . |
| AMC38 | HC | M | 51 – 55 | NO | . | . | . | 0 | . | . | . | . | . | X |
| AMP33 | PrEP | M | 26 – 30 | NO | Truvada | Emtricitabine, Tenofovir | . | 0.1 | X | X | X | . | X | . |
| AMP35 | PrEP | M | 21 – 25 | YES | Truvada | Emtricitabine, Tenofovir | . | 1.0 | X | X | X | X | X | . |
| AMP32 | PrEP | M | 35 – 40 | NO | Truvada | Emtricitabine, Tenofovir | . | 1.1 | X | X | . | . | . | . |
| AMP23 | PrEP | M | 61 – 65 | YES | Truvada | Emtricitabine, Tenofovir | . | 1.3 | X | . | . | . | . | . |
| AMP30 | PrEP | M | 26 – 30 | NO | Truvada | Emtricitabine, Tenofovir | . | 2.4 | X | . | . | . | . | . |
| AMP31 | PrEP | M | 35 – 40 | NO | Truvada | Emtricitabine, Tenofovir | . | 2.5 | X | X | . | . | . | . |
| AMP34 | PrEP | M | 41 – 45 | NO | Truvada | Emtricitabine, Tenofovir | . | 3.2 | X | X | X | . | . | . |
| AMP36 | PrEP | M | 26 – 30 | YES | Truvada | Emtricitabine, Tenofovir | . | 3.4 | X | X | X | X | X | . |
| AMP27 | PrEP | M | 35 – 40 | NO | Truvada | Emtricitabine, Tenofovir | . | 4.4 | X | X | X | X | X | . |
| AMP32.2<sup>c</sup> | PrEP | M | 35 – 40 | NO | Truvada | Emtricitabine, Tenofovir | . | 4.3 | . | . | X | X | X | . |
| AMP32.3<sup>c</sup> | PrEP | M | 41 – 45 | NO | Truvada | Emtricitabine, Tenofovir | . | 5.2 | . | . | . | . | . | X |
| AMP29 | PrEP | M | 35 – 40 | YES | Truvada | Emtricitabine, Tenofovir | . | 5.4 | . | X | . | . | . | . |
| AMP23.2 ° | PrEP | M | 66 – 70 | YES | Truvada | Emtricitabine, Tenofovir | . | 5.9 | . | . | . | . | . | X |
| AMP36.2 ° | PrEP | M | 31 – 35 | YES | Truvada | Emtricitabine, Tenofovir | . | 6.5 | . | . | . | . | . | X |
| AMV23 | PWH | M | 31 – 35 | YES | Genvoya | Emtricitabine, Tenofovir, Cobicistat, Elvitegravir | 0.7 | 0.6 | X | X | X | X | X | . |
| AMV27 | PWH | M | 56 – 60 | NO | Descovy, Isentress | Emtricitabine, Tenofovir, Raltegravir | 1.1 | 1.0 | X | X | . | . | . | . |
| AMV17 | PWH | M | 41 – 45 | YES | Viread, Tivicay, Edurant | Tenofovir, Dolutegravir, Rilpivirine | 3.7 | 2.7 | X | X | X | X | X | . |
| AMV23.2 ° | PWH | M | 35 – 40 | YES | Genvoya | Emtricitabine, Tenofovir, Cobicistat, Elvitegravir | . | . | . | . | . | . | . | X |
| AMV21 | PWH | M | 61 – 65 | YES | Truvada, Tivicay | Emtricitabine, Tenofovir, Dolutegravir | 6.2 | 5.7 | X | X | X | X | X | . |
| AMV5 | PWH | M | 56 – 60 | NO | Truvada | Emtricitabine, Tenofovir | 21.5 | 8.5 | X | X | . | . | . | . |
| AMV26 | PWH | M | 46 – 50 | NO | Atripla | Emtricitabine, Tenofovir, Efavirenz | 18.4 | 8.5 | X | X | . | . | . | . |
| AMV25 | PWH | M | 56 – 60 | NO | Atripla | Emtricitabine, Tenofovir, Efavirenz | 11.6 | 11.3 | . | X | X | . | X | . |
| AMV19 | PWH | M | 56 – 60 | YES | Triumeq | Abacavir, Dolutegravir, Lamivudine | 12.7 | 11.4 | X | X | . | . | . | . |
| AMV15 | PWH | M | 51 – 55 | NO | Triumeq | Abacavir, Dolutegravir, Lamivudine | 14.1 | 13.7 | X | X | X | X | X | . |
| AMV9 | PWH | M | 56 – 60 | YES | Truvada, Isentress | Emtricitabine, Tenofovir, Raltegravir | 14.8 | 13.8 | X | X | X | . | . | . |
| AMV16 | PWH | M | 56 – 60 | YES | Genvoya | Emtricitabine, Tenofovir, Cobicistat, Elvitegravir | 30.1 | 15.7 | X | . | X | X | X | . |
| AMV4 | PWH | M | 56 – 60 | YES | Biktarvy | Emtricitabine, Tenofovir, Bictegravir | 17.0 | 17.0 | X | X | X | . | . | . |
| AMV11 | PWH | M | 46 – 50 | YES | Complera | Emtricitabine, Tenofovir, Rilpivirine | 19.9 | 18.9 | X | X | X | X | X | . |
| AMV3 | PWH | M | 51 – 55 | NO | Truvada, Isentress | Emtricitabine, Tenofovir, Raltegravir | 21.4 | 19.4 | X | X | . | . | . | . |
| AMV14 | PWH | M | 56 – 60 | YES | Kivexa, Reyataz, Norvir | Abacavir, Lamivudine, Atazanavir, Ritonavir | 21.2 | 20.5 | X | X | X | X | . | . |
| AMV20 | PWH | M | 56 – 60 | YES | Truvada, Tybost, Prezista | Emtricitabine, Tenofovir, Cobicistat, Darunavir | 21.3 | 20.7 | X | X | . | . | . | . |
| AMV10 | PWH | M | 56 – 60 | NO | Intelence, Prezista, Tivicay, Norvir | Etravirine, Darunavir, Dolutegravir, Ritonavir | 25.4 | 21.5 | X | X | . | . | . | . |
| AMV13 | PWH | F | 51 – 55 | YES | Complera, Reyataz, Isentress | Emtricitabine, Tenofovir, Rilpivirine, Atazanavir, Raltegravir | 23.1 | 23.1 | X | X | X | X | X | . |
| AMV11.2 ° | PWH | M | 56 – 60 | YES | Complera | Emtricitabine, Tenofovir, Rilpivirine | 25.8 | 24.2 | . | . | . | . | . | X |
| AMV3.2 ° | PWH | M | 56 – 60 | NO | Truvada, Isentress | Emtricitabine, Tenofovir, Raltegravir | 27.1 | 25.1 | . | . | . | . | . | X |

<sup>a</sup> Cigarettes and/or cannabis. <sup>b</sup> Fraction of years from the ART start to BAL collection. <sup>c</sup> BALs of the same subject collected at different dates. ART, antiretroviral therapy; BAL, bronchoalveolar lavage, HC, healthy controls; PrEP healthy subject on pre-exposure prophylaxis; PWH, people with HIV under ART; an X indicate samples employed in each of the five tested assays.

To assess the epigenetic and transcriptomic state of AM we used five different assays (**Fig. 1a**). We tested the hypothesis that ART imprinted a physiological state in AM that primed pathways linked to pulmonary diseases. We carried out the bulk testing in three separate steps (**Fig. 1b**). (i) We compared the epigenetic and transcriptomic state of AM between the HC, PrEP and PWH groups independent of time on ART. (ii) We tested the effect of time on ART in PrEP and PWH individuals separately. Accessibility of chromatin regions, histone marks, and gene expression changes with increased time for both groups were denoted as having an ART effect. (iii) We conditioned the effect of continuous exposure to ART on the PrEP/PWH/HC group status to highlight the main biological mechanisms affected by continuous exposure to ART. Next, we employed snMulti to assess AM subpopulation specific effects on models (i) and (iii); and (iv) to evaluate group differences to SARS-CoV-2 challenge, which we used as an experimental probe for the AM immune response (**Fig. 1b**).

### Bronchoalveolar lavages

Bronchoscopies were performed at the Centre for Innovative Medicine of the MUHC as per American Thoracic Society guidelines and as previously described (17, 23). Briefly, standard flexible bronchoscopy of the middle lobe was conducted under local anesthesia with lidocaine with additional conscious sedation with intravenous midazolam. A total of 200 ml sterile saline instilled in 50-ml volumes resulted in an average return of 115 ml of BAL fluid. BAL fluid in non-adherent tubes was stored on ice for a maximum of 30 minutes. Prior to further processing, BAL fluids were strained through 100μm filter to remove any mucus and cell clumps. Cytology assessment was performed for all BAL samples and indicated that AMs represented >85% of all recovered mononuclear cells. Collected BAL cells were spun for 10 minutes at 300g, 10°C, and washed twice in RPMI-1640 with L-glutamine (Wisent, Canada), containing 2.5% human serum (heat-inactivated AB+ off the clot, Wisent, Canada), 1% penicillin/streptomycin (Gibco, USA), 10mM HEPES (Gibco, USA), 1% non-essential amino acids (Gibco, USA) and 2.5 µg/ml Amphotericin B (Wisent, Canada). BAL cells were counted and seeded at ∼250K cells/well in low attachment 24-well plates (Nunclone Sphera®) for culture and viral infection.

### Library preparation

Data for bulk RNAseq, ATACseq, and H3K27ac was generated previously and the methodology used to prepare libraries and to process the data for RNA-, ATAC- and H3K27ac was described in (17). For this study, we added two histone marks: H3K4me3 and H3K27me3. We also expanded the dataset for the H3K27ac mark. Libraries for the three histone marks were produced using ChIPmentation kits for histones from Diagenode (C010110009). Aliquots of crosslinked AM containing 10^5^ cells stored at -80°C were thawed and resuspended on 50 µL of ice-cold Sarkosyl buffer. Next, samples were sonicated on Bioruptor Pico to obtain fragments between 200 – 500 bp. ChIPmentation was carried out using the automated IP-Star Compact with Diagenode recommended protocol and the Diagenode antibodies for H3K4me3 (C15410003), H3K27ac (C15410196), and H3K27me3 (C15410195). After overnight incubation, DNA fragments captured by the immunoprecipitation were reverse crosslinked and unique barcodes, Diagenode (C01011034, C01011036, and C01011037), were added to the sequences of each subject and histone mark. Finally, libraries were sequenced on Illumina NovaSeq 6000 S4 PE100 aiming for an average of 50 million reads per library. The raw sequences were then processed using the same protocol previously reported (17).

For the snMulti, nuclei preparation was done following an adapted 10xGenomics protocol (CG000365). Specifically, to obtain good quality AM nuclei from BAL cells were lysed while beating with 3mm glass beads for 4 second on MP Biomedicals™ FastPrep-24™ (speed 6.5). Next, ATAC and gene expression libraries were constructed following manufacturer’s instructions using 10xGenomics Epi multiome ATAC + Gene Expression kit (CG000338). snMulti libraries were paired-end sequenced on Illumina NovaSeq 6000 S4 flow cells.

### snMulti quality control and data integration

Single-nucleus multimodal sequencing data was processed using 10X cellranger-arc v.2.0.2 with default parameters. Barcoded reads from each single nucleus were aligned to the human genome GRCh38. Data processing was performed per modality prior to multimodal integration. First, HDF5 files with gene expression count were imported into R using BPCells v.0.3.0 (24) and processed with Seurat v5.2.1 (25). Nucleus with mitochondrial reads > 0.2 and detectable genes < 300 were excluded. Next, chromatin accessibility counts and fragments for nucleus that passed quality-control for the RNA modality were imported to Seurat and processed using Signac v.1.14.0 (26). We used a unified peak set across all samples to recalculate feature matrix counts before snATAC integration. We then estimated snATAC parameters and excluded nucleus with fraction of reads in peaks < 0.6, nucleosome signal < 1.5, TSS enrichment < 1, and counts < 1000.

The snRNA modality was integrated using the standard Seurat multiomics pipeline. The data for each library was normalized with SCTransform, and integration was performed using *IntegrateLayers* with reciprocal PCA (RPCA) using one unchallenged library from each group (HC, PrEP, and PWH) as a reference and the top 30 PCs. Dimensionality reduction and clustering were performed using *FindNeighbors, FindClusters*, and *RunUMAP*, to define cells subpopulations and to separate AM from DCs and lymphocytes, which represented between 0.1% and 10% of total cells from BAL. snATAC integration followed Signac pipeline. Term frequency–inverse document frequency (TF-IDF) normalization was applied, followed by latent semantic indexing (LSI) via *RunSVD*. The LSI 2 to 15 were used for dimension reduction and clustering. The first LSI component was excluded from the integration as it captures sequencing depth. Cell subpopulations were identified using the same approach as for the snRNA modality. Finally, we performed weighted nearest neighbor (WNN) analysis using Seurat *FindMultiModalNeighbors* to integrate the RPCA and LSI reductions from snRNA and snATAC, respectively. snMulti WNN clusters were used to define the AM subpopulations shown throughout the manuscript.

### Single nucleus annotation

To identify the main biological processes characterizing the physiological state of AM subpopulations we used Seurat *FindConservedMarkers* for unchallenged, and SARS-CoV-2 challenged libraries separately. Next, we used cluster markers with positive fold changes expressed in > 25% of the unchallenged AM for each subpopulation to perform a gene ontology (GO) analysis using *enrichGO* form clusterProfiler (27). We selected the most significant GO term per subpopulation as a proxy of the main physiological state of the AM clusters. We then calculated a module score using *AddModuleScore* using the cluster marker genes per nucleus for the top GO terms and indicated the density of the biological state in the UMAP using *plot_density* from Nebulosa v.1.0.1 (28). To identify a transitional state of the AM subpopulations we performed pseudotime and trajectory analysis using unchallenged libraries with slingshot v.2.14.0 (29).

### snMulti SARS-CoV-2 ex vivo challenge

We used the SARS-CoV-2 strain RIM-1 isolated from a patient in Quebec (GenBank ID MW5997360) (30). The SARS-CoV-2 strain was propagated using VeroE6 cells infected at a multiplicity of infection (MOI) of 0.05 TCID50/cell and incubated in DMEM at 37 °C for 3 days. The virus-containing supernatant was harvested, clarified by centrifugation at 2,000g for 5 min. Cell-associated viruses were obtained by 3 freeze/thaw cycles, where cell debris was removed by centrifugation at 4,000g. Viral stocks were concentrated using Amicon® Ultra-15 100K Centrifugal Filter Units. BAL cells were *ex vivo* challenged with SARS-Cov2 at MOI 2:1 TCID50/cell or left untreated overnight (16-18 hours) in culture medium RPMI (Gibco) with 2.5% human serum, 1% penicillin/streptomycin, 10mM HEPES, 1% non-essential amino acids and 2.5 µg/ml Amphotericin B. Further, cells were collected by gentle pipetting, washed twice in PBS (Wisent), 0.04% BSA (Wisent) and 20 µg/ml DNase I (Roche) to prevent cell clumping and assessed for viability.

### Statistical modeling

Bulk differences in epigenetics and transcriptomics of AM were tested with three models comparing: (i) groups: PrEP, PWH and HC; (ii) time on ART for PrEP and for PWH, and (iii) continuous exposure to ART adjusted by group with the following linear modeling in Limma v3.60.2 (31):

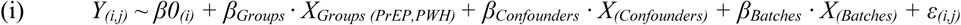

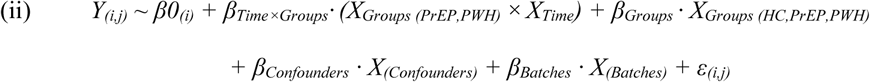

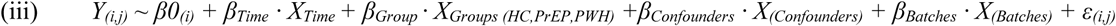

Where *Y*_(i,j)_ represents the estimate quantification for each feature “i” and sample “j”. Briefly, quantification matrices with *Y*_(i,j)_ were depth-normalized with *calcNormFactors* upperquartile (0.75) for the epigenetic assays and TMM for the transcriptomic data using edgeR v.3.40.2. The mean-variance relationship and log2 count per million was estimated with *voom* v3.60.2 (32). In these models, *β0_(i)_* represents the intercept for the feature “i”. In model (i) the intercept corresponds to the HC group. In models (ii) and (iii) the intercept represents the predicted value of *Y* when the continuous variables of interest (e.g. Time on ART) are equal to zero. In model (i) *β_Groups_* output two *β* that captures the main effect for PrEP and PWH relative to the intercept (HC), respectively. The contrast between PrEP and PWH was done post-hoc by contrasting their respective *β* coefficients. In models (ii) and (iii) the *β_Groups_* adjusts for the main effect of group membership on the “Time” effect. In model (ii) the *β_Time×Groups_* is an interaction term that output two *β* that captures the effect of time on ART for PrEP and for PWH. In model (iii) the *β_Time_* term estimates the effect of continuous exposure to ART adjusting for the main group effect. The coefficients of *β_Confounders_* included the phenotypic confounders: sex, chronological age, smoking status, *Mtb* challenge, and a seasonal effect of BAL collection. The coefficients of *β_Batches_*included the technical covariates: sequencing batch, fractions of reads in peaks for the epigenetic assays, time of AM incubation, and number of AM seeded per plate. The *ε_(i,j)_* represents the residual error not explained by variables in the model. A Benjamini–Hochberg (BH) procedure was used to estimate False Discovery Rate (FDR) for each assay in the three tested models. Genes and peaks presenting FDR < 10% were selected for further follow-up on the pathway and transcription factors analyses.

snMulti differential analysis was performed using a pseudo bulk approach per AM subpopulations. Briefly, pseudo counts per AM subpopulations, subject, and challenge status were calculated using Seurat *AggregateExpression*. Next, genes expressed in less than 20% of the nuclei of a subpopulation for the snRNA modality and peaks quantified in less than 5% of the snATAC modality were excluded from the analysis. For the snATAC pseudo bulk we filtered for peaks linked to genes. Chromatin region from snATAC were assigned to genes using Signac *LinkPeaks*. Peaks that correlated with flanking gene expression within 250kb with a Pearson correlation > 0.01 and *p* value < 0.1 were selected for downstream analysis. We used this lenient cut off to avoid removing epigenetic features that lacked stronger correlations with genes due to the sparsity of the snATAC data. The pseudo counts per modality was performed as in bulk (32, 33). The statistical analyses testing differences between groups and continuous exposure to ART applied the same statistical framework of models (i) and (iii). To test AM response to SARS-CoV-2 challenge per group we used a per subject paired design as indicated in the equation below.

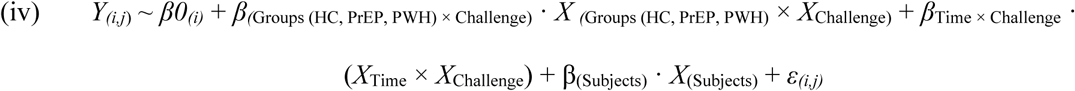

In this model, β0*_(i)_* represents the intercept which corresponds to one unchallenged HC library followed by two interaction terms. The first interaction term captures the effect of SARS-CoV-2 challenge with one output *β* per Group (HC, PrEP, and PWH) and the second interaction term *β* captures the effect of SARS-CoV-2 according to the time of continuous exposure to ART. Then one *β* per subject characterizes the blocking design and the *ε_(i,j)_* represents the residual error not explained by variables in the model.

### Bulk peak to gene assignment and Gene-Set Co-Regulation Analysis

Genes were assigned to peaks using *annotatePeak* from ChIPSeeker v.1.40 (34). Each peak from the epigenetic assays was assigned to the nearest gene and to any additional flanking gene at < 5 kilobases from a peak. The gene set co-regulation analysis (GESECA) was performed with *geseca* from *fgsea* v1.30 (35). Briefly, GESECA identifies patterns of co-expression, co-accessibility or co-marks enriched within predefined gene sets across samples. GESECA was developed to compare datasets with a large number of different biological conditions such as timepoints (e.g. time on ART). It uses as input a quantification or a covariate-reduced matrix (gene/peak by samples) to calculate the proportion of variance between samples explained by each gene set. *P*-value significance for each gene set is then calculated by permutation of genes between lists. For the GESECA we used covariate-regressed matrices obtained via *removeBatchEffect* from *Limma* for transcriptome and epigenome. By design, GESECA does not directly compare phenotypic groups but rather variability between samples (i.e., individuals GESECA scores). To assess if the variability between samples was driven by group or a time effect, we contrasted the GESECA score per subject with the phenotype of interest in each model. For group comparisons, we used a Wilcoxon test to estimate significant differences between the median GESECA score per group. To test the correlation of time and GESECA scores we used linear regression both for the per-group time comparisons (PrEP and PWH) and the overall effect of continuous exposure to ART. An FDR correction for these comparisons was calculated using BH. We ran GESECA using the HALLMARK pathway gene-set collection from MSigDB (36). Additionally, as senescence is a hypothesized mechanism contributing to comorbidities in the aging PWH we included the REACTOME “Senescence” branch, and SenMayo, a well-established senescence gene set across tissues and species in the gene-set lists (37, 38).

### Motif enrichment analysis

To test for enrichment of transcription factor binding motifs in differential regions associated with the continuous exposure to ART, we used *findMotifsGenome* from HOMER v4.11 (39). Differential accessible chromatin regions (ATAC) were compared to (i) 500,000 random regions matched by size and GC content and (ii) nonsignificant 20,060 regions at FDR > 50% in the continuous exposure to ART analysis. We considered a motif to be significantly enriched if it presented FDR < 1%. Pairwise correlation of TF motif binding by similarities was performed with STAMP v.1.3 default parameters (40).

### Footprint analysis

Transcription factor Occupancy also known as Footprint analysis was performed with TOBIAS v0.14.0 using covariate-regressed ATAC traces (41). Briefly, we adapted the concept of regressing out known confounders (e.g. covariables and batch effects) prior to chromatin modeling as proposed in (42, 43). First, signal intensity for regions with increased chromatin accessibility (ATAC) with continuous exposure to ART was calculated from BAM files of each subject using *bamCoverage* from deepTools v.3.5.1 with --binSize 1 and --normalizeUsing CPM (44). Next, ATAC signals per sample were summarized using *multiBigwigSummary* (44). The output of this procedure was a quantification matrix with ATAC signal intensity at 1bp resolution per subject. This matrix was then exported into R v4.4.0. We used *removeBatchEffect* from *Limma* to calculate the residual signal intensity after regressing the coefficients *β*_Group (HC,PrEP,PWH)_, *β*_Confounders_ and *β*_Batch_ while preserving *β_0_* and *β_Time._* from the model (iii). This residual signal intensity matrix represents ATAC traces mimicking the linear model (iii) tested in the differential analysis. The residual traces were exported from R and converted back to individual BigWigs using *bedGraphToBigWig* from UCSC. Next, we used TOBIAS *ScoreBigwig* to identify regions of protein binding across differential regions with continuous exposure to ART. To predict which TF bind to the footprints detected, we combined footprint scores per subject with information on TF binding motifs using TOBIAS *BINDetect*. We used as reference 919 motifs from the JASPAR CORE database for vertebrates (45). *BINDetect* calculates a mean footprint score for each of the 919 TF binding sites split per subject. This score can be compared between samples to access differential changes as TF having high-scoring footprints denotes a clearer and more defined bound. To test if the TF footprint intensity was associated with time, we fit a linear model comparing the footprint mean intensity per TF with the continuous exposure to ART. Fold change per year was calculated via the exponential of the model slopes and FDR *p*-values were adjusted with BH. TOBIAS *PlotAggregate* was used to summarize traces for selected examples of TF. The residual signal intensities for selected examples of footprints were plotted per timepoint and according to their chromosomal position for each assay using *ggplot2* v.3.4.4 with *geom*_*spline* from *ggformula* v.0.10.4 (46, 47).

### snMulti transcription factor signature and gene regulatory network

We infer TF activity signature using decoupleR v2.4.1 as previously described (48, 49). Briefly, TF – target gene interaction network was obtained from the collecTRI database. We filtered the snRNA modality for genes significantly associated with continuous exposure to ART (FDR < 10%) within the AM subpopulation. TF activity scores were computed per cell using *run_ulm*, which fits a linear model that predicts the observed gene expression based solely on the TF-Gene interaction weights, with the t-value of the slope used as the activity score. To evaluate the relationship between TF activity and continuous exposure to ART, we modeled TF scores as a function of time, adjusting for group (HC, PrEP, PWH) using the *lm* function in base R. *β* coefficients representing the effect of continuous exposure to ART on TF activity were extracted, and a BH FDR was calculated. TFs showing absolute *β* > 0.02 and FDR < 0.01 were considered significant.

To infer the multimodal TF gene regulatory networks (GRN) associated with continuous exposure to ART, we applied the Pando v1.0.2 framework (50). Pando models gene expression as a function of the interaction between TF expression and chromatin accessibility at regions with TF binding sites per nucleus. For the GRN analysis, we tested 627 TFs from the JASPAR2020 human CORE collection (51). As candidates for the GRN, we evaluated all differential features for snRNA (n = 867) and snATAC (n = 3,332) associated with continuous exposure to ART. Regulatory TF interactions were modeled with *infer_grn* using the Signac peak-to-gene method, a 250 kb upstream and 50 kb downstream window, and multiple testing correction using the BH procedure. Modules of co-regulated genes were identified using *find_modules*, and modules with FDR < 10% were considered significant.

## Results

### Differences in AM physiology between PWH, people on PrEP and HC

To assess the correlation of epigenetic and transcriptomic features (i.e., accessible chromatin, H3K27me3, H3K27ac, H3K4me3, and RNA) with study group we performed a principal component analysis (PCA). After accounting for known confounders (smoking, sex, chronological age, and batch) and preserving the phenotype group variable, the top four principal components (PC) captured between 38% to 75% of the remaining variability. Yet, the top components only partially clustered subjects according to their respective groups suggesting strong interindividual variability (**Fig. 2a**). As a result, when performing the differential feature analysis, only a small proportion of the regions tested passed with FDR < 10% while differences in gene expression were more pronounced (**Fig. S1; Table S1**). Among genes with decreased expression in PrEP, we identified all 13 mitochondrial-encoded genes of the electron transport chain (**Fig. S2**). This indicated a potential mitochondrial dysfunction in AM from PrEP that was also observed to a lesser extent for PWH.

**Fig. 2.**
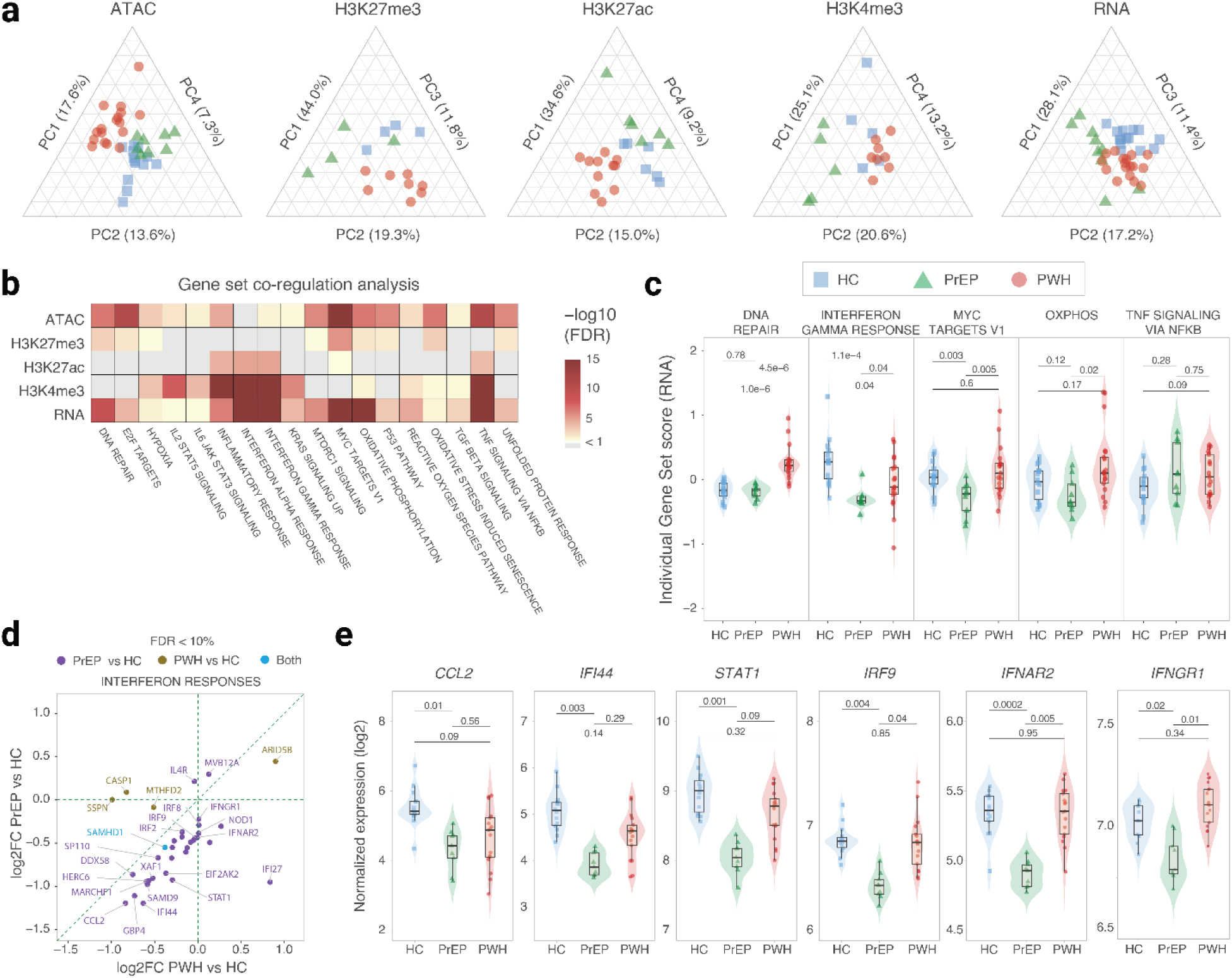
Epigenetic and transcriptomic differences between AM from people on PrEP, PWH and HC. **(a)** Phenotypic group clustering using the top principal components (PCs). Scaled PCs and their corresponding percentage of variance explained are shown on each axis of the ternary plots for the five tested assays. **(b)** Gene Set Co-regulation Analysis (GESECA) for Hallmark pathways. Gene sets with False Discovery Rate (FDR) < 10% in at least three assays. Results for all tested pathways are given in Table S2. (**c**) GESECA scores for the top five differential pathways. The scores of the transcriptomic assay for each subject are indicated on the y-axis separated by group on the x axis. (**d**) Correlation between differential expression for interferon response genes in PrEP and PWH versus HC. The log2 fold difference (log2FC) between PrEP and HC was plotted against the log2FC for PWH vs HC for the transcriptomic assay. Each circle represents a gene in the Hallmark interferon pathways that had FDR < 10%. **(e)** Normalized gene expression per subject and group for selected genes in HALLMARK interferon pathways. In panels “c” and “e”, a Wilcoxon test was used to estimate significant differences in the median between groups with the FDR adjusted *p*-values shown at the top for each group comparison.

Given the pronounced interindividual variability across all studied groups, we performed a GESECA to identify pathways associated with the most highly variable features (see methods). Using this approach, we found that differences between subjects were significantly enriched in inflammatory, cellular homeostasis and stress related pathways both at the transcriptomic and epigenetic levels (**Fig. 2b; Table S2**). Next, we evaluated if the differential pathways between subjects were driven by the phenotypic groups (**Fig. 2c**). We found that genes assigned to the DNA repair pathway had significantly increased expression in AM from PWH compared to PrEP and HC. DNA repair is a known mechanism activated in host response to HIV integration. In contrast, expression of genes assigned to interferon pathways were significantly depleted in AM from PrEP and PWH compared to HC (**Fig. 2c**). Of the 183 genes of hallmark interferon pathways that are expressed in AM, 28 (15.3%) had decreased expression in PrEP compared to HC with most of these genes displaying a trend of decreased expression in PWH (**Fig. 2d**). Among key genes with lower expression in PrEP were Interferon-Inducible genes (IFIs), Interferon Regulatory Factors (IRFs), and interferon receptors both type-I (*IFNAR2*) and type-II (*IFNGR1*) (**Fig. 2e; Table S1**).

Overall, the group comparison indicated an active DNA damage repair (DDR) mechanism in AM from PWH and a constitutive lower expression of interferon-related genes in AM that was more pronounced in PrEP participants. On the other hand, the inter-individual variabilities associated with TNF signaling pathways were not captured by group assignment (**Fig. 2c**). The lack of association between gene set scores for these pathways and group labels suggested that unaccounted participant characteristics were likely driving the inter-individual variability. However, when displaying the time of continuous exposure to ART for the individuals in Fig. 2c we observed a strong trend for a time dependent effect on TNF signaling for AM from both PrEP and PWH (**Fig. S3**). We therefore hypothesized that time on ART for PrEP and for PWH is an important factor to explain differences in AM state between subjects.

### The effect of time on ART on the transcriptomic and epigenetic landscape of AM

While accounting for the same covariates as in the previous analyses, we added a time variable to the model and tested PrEP and PWH groups independently. The time variable captured the fraction of years from the start of the ART regimen to BAL collection (**Table 1**). To avoid regressing out the variable of interest, we conducted a PCA on the residuals after removing confounders and preserving the time variable. With this approach we showed that PC1 strongly correlated with time on ART in both PrEP and PWH in all tested assays (**Fig. 3a**). We observed a high proportion of significant differential epigenetic and transcriptomic features at FDR < 10% for time on ART for PrEP and length of time since ART initiation for PWH (**Fig. 3b; Table S3**). Chromatin accessibility and the H3K27ac mark showed the highest numbers of significantly differential features affected by time in both groups. Additionally, PWH presented >5-fold more time-dependent differentially expressed genes compared to the PrEP participants and more active promoters with increased H3K4me3 (**Fig. 3b; Table S3**). This may reflect HIV-specific effects, time-dependent changes not detected in the PrEP group due to the shorter ART regimen, or higher statistical power due to the larger sample size of the PWH group. To compare features affected by time on ART in both groups, we intersected genes and features significantly associated with time in the PrEP and PWH groups (**Fig. 3c**). With the exception of the promoter repressor H3K4me3 mark, we identified a significant enrichment of changes occurring in the same direction for time on ART in both PrEP and PWH groups (**Fig. 3c**).

**Fig. 3.**
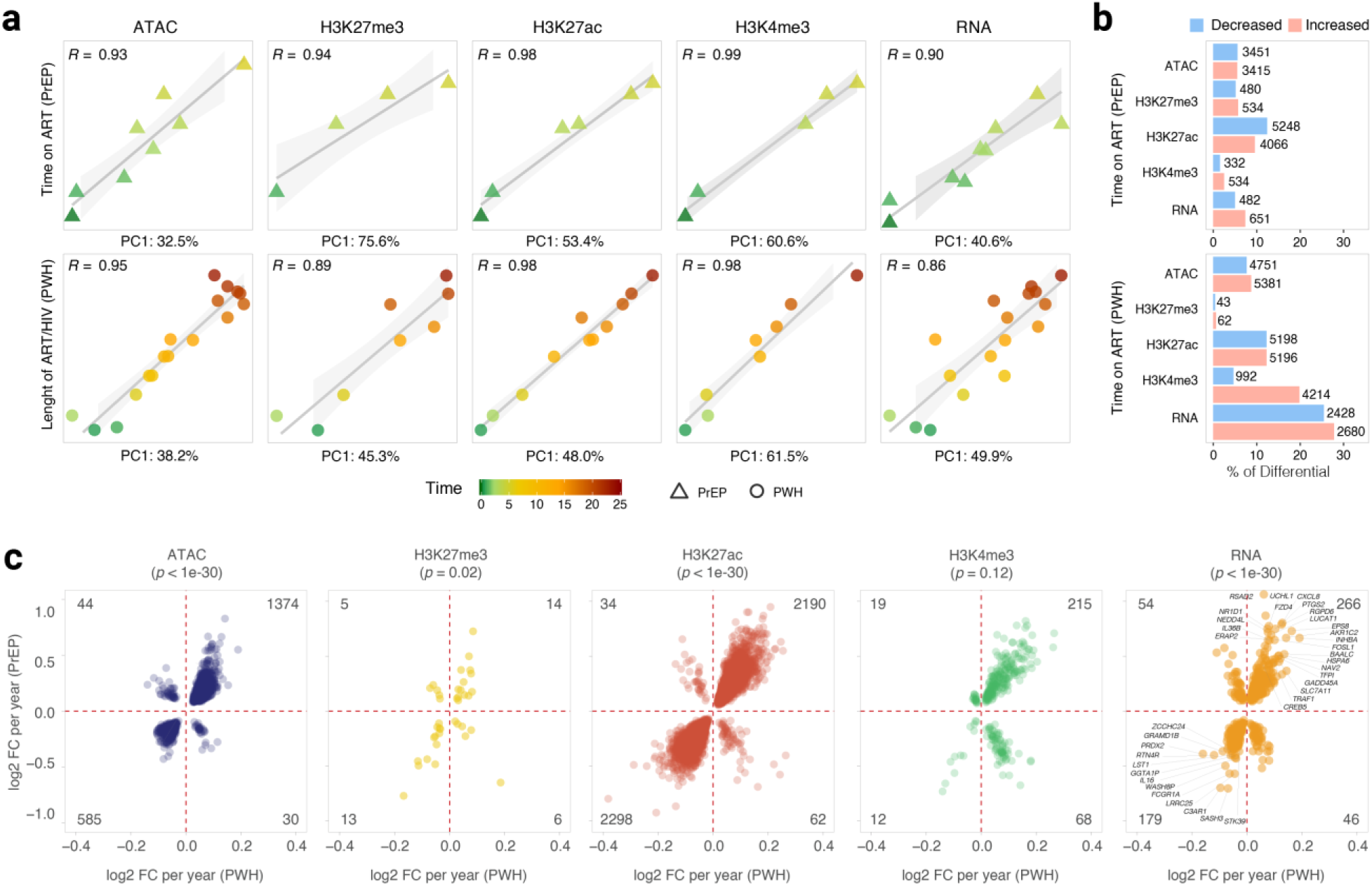
Differential epigenetic and transcriptomic features with time on ART for PrEP and PWH. (**a**) The proportion of variance captured by the first principal components was plotted against time on ART for people on PrEP (top) and for PWH (bottom). The Pearson correlation R is shown at the left upper corner of each plot. (**b**) Epigenetic and transcriptomic features significantly associated with time on ART in PrEP and PWH participants. The bar plots show the proportion of differential regions/genes at an FDR of 10% relative to the total number of tested regions/genes for time on ART in PrEP (top) and in PWH (bottom). The absolute numbers of differential regions/genes are shown next to the bars. Decreased and increased DNA accessibility, histone marks, and gene expressions are shown in blue and red, respectively. (**c**) Epigenetic and transcriptomic features significantly changed in the same direction with time for PrEP and PWH participants. Only features with FDR < 10% in both groups are plotted

### Biological mechanisms affected by time on ART in PrEP and PWH

To assess the biological mechanisms affected by time on ART we performed a GESECA analysis separately for the PrEP and PWH groups (**Table S2**). We highlighted 15 gene sets with the highest variability between samples within the PrEP and PWH groups (**Fig. 4a**). These terms included OXPHOS, immune mediators, control of cellular stress, cell cycle arrest, and senescence (**Fig. 4a**). Next, we tested if the per individual GESECA score of highlighted pathways correlated with time on ART in PrEP and PWH (**Fig. 4b; Fig. S4**). At the RNA level, 14 out of 15 pathways significantly correlated with time in at least one group (*P* ≤ 0.01) (**Fig. 4b; Fig. S4**). At the epigenetic level, time on ART correlated (*P* ≤ 0.01) with individual GESECA scores in all highlighted pathways for at least one active epigenetic assay (ATAC, H3K27ac, and H3K4me3). Decreased expression of genes assigned to the OXPHOS pathway correlated with time on ART in PWH, but not in the PrEP group (**Fig. 4b**). However, hallmark OXPHOS pathway do not encompass mitochondrial genes of the electron transport chain that had decreased gene expression in PrEP and PWH independent of time on ART (**Fig. S2**). This suggested that time on ART may impact different components of OXPHOS at different levels. This is supported by the decreased expression of autosomal genes encoding members of the electron transport chain complexes I–V that was more pronounced with increased time on ART in PWH (**Fig. 4c**).

**Fig. 4.**
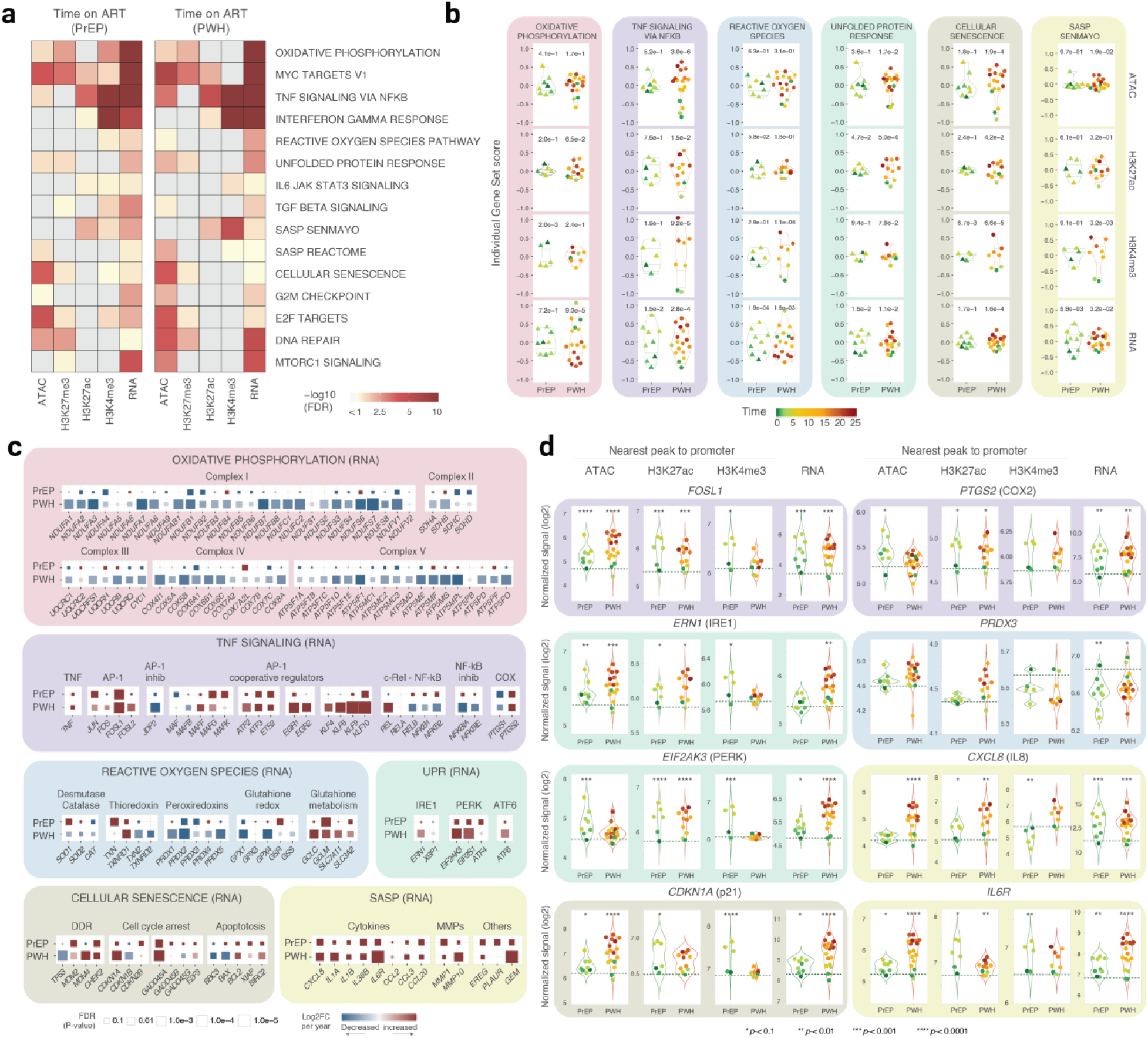
Biological pathways affected by time on ART in people on PrEP and in PWH. (**a**) GESECA gene sets with the largest variability between participants within each group. Terms with FDR < 10% in at least two assays in each group are shown as a heatmap. Data for all tested gene sets is given in Table S2. (**b**) Per subject GESECA scores (y-axis), separated by group (x-axis) for six gene sets affected by time on ART. Remaining pathways are shown in Fig. S4. The quantiles are shown as violins for reference of the distribution. A linear model was used to test the correlation of time on ART in PrEP and PWH with the individual GESECA scores for all tested assays. *p-*values for the correlations are shown at the top of each comparison. The repressor mark H3K27me3 did not show a significant correlation with time in any group and was omitted. (**c**) Changes in gene expression for selected genes belonging to pathways shown in panel “b”. Genes were grouped by main function in each pathway. The absolute values for log2FC and FDR are shown in Table S3. (**d**) Promoter accessibility and histone marks, and expression of selected genes affected by time on ART in both groups. A dotted horizontal green line indicates the average signal in healthy controls for reference. *p-*values are given at the top of the group for each comparison with the absolute values and FDR shown in Table S4.

Transcription of genes assigned to the TNF signaling pathway correlated with time on ART in both the PrEP and PWH groups (**Fig. 4b**). The increase in the transcription-based GESECA score per year for the TNF pathway was higher in PrEP compared to PWH (**Fig. 4b**). Among the genes assigned to TNF signaling that displayed increased expression with time on ART were transcription factors that are key mediators of immune responses and senescence (**Fig. 4c**). For example, subunits of the AP-1 complex (e.g. *JUN* and *FOSL1*), and AP-1 cooperative regulators (e.g. *EGR1*, *ATF3*, *ETS2,* and *KLF10*) had increased expression with time on ART in AM from subjects on PrEP and PWH (**Fig. 4c**). Moreover, key pro-inflammatory mediators such as *PTGS2* (i.e. COX-2) and *TNF* showed increased expression with time on ART in both groups, albeit *TNF per se* did not surpass FDR correction (**Fig. 4c**). For these key genes we also identified a time-related effect at the epigenetic level. For example, the transcription starting site (TSS) of *FOSL1* and *PTGS2* had increased chromatin accessibility, as well as H3K27ac and H3K4me3 marks with increased time on ART (**Fig. 4d**).

Time on ART also impacted on the AM cellular response to stress via inactivation of reactive oxygen species (ROS) and protein turnover via the unfolded protein response (UPR) (**Fig. 4b**). For the ROS pathway we noted a distinct effect of time for AM from the PrEP and PWH groups at the RNA level (**Fig. 4b**). However, this was due to groups capturing different aspects of the ROS pathway. For example, redoxins (e.g. PRDXs and GPXs) had more decreased expression with time on ART in PWH with a less pronounced effect in PrEP (**Fig. 4c**). Conversely, glutathione metabolism substrates used by GPXs to neutralize free radicals, and glutathione reductase (*GSR*) had increased expression with time on ART in PrEP (**Fig. 4c**). Overall, AM presented an unbalanced expression between effector enzymes and substrate metabolism when coping with cellular stress. The UPR pathway plays a crucial role in maintaining cellular homeostasis and encompasses three main branches via IRE1, PERK and ATF6. Notably, the *ERN1* (IRE1), *EIF2AK3* (PERK) and *ATF6* genes had increased expression with time on ART in PWH (**Fig. 4c**). The PERK branch genes also had increased expression with time on ART in PrEP (**Fig. 4c**). Interestingly, at the epigenetic level, the promoters of *ERN1* and *EIF2AK3* were more active with increased time on ART in both PrEP and PWH groups (**Fig. 4d**). Taken together, our data suggested a dysregulated response to stress in AM with increased time on ART.

Accelerated cellular aging has been suggested as a candidate mechanism contributing to the increased incidence of comorbidities in PWH. We found that the Cellular Senescence and Senescence Associated Secretory Phenotype (SASP) gene sets correlated with time on ART (**Fig. 4b**). Senescence is a complex mechanism that involves multiple cellular processes including DDR, cell cycle arrest, cellular stress and cytokines mediating the immune response. Increased AP-1 activity, and levels of p21 (encoded by *CDKN1A*), as well as Growth Arrest and DNA Damage-inducible genes (i.e. *GADD45A*) are considered a hallmark of cellular senescence via cell cycle arrest (**Fig. 4c**). The *CDKN1A* promoter was progressively more active with increased chromatin accessibility, H3K27ac and H3K4me3 marks, and *CDKN1A* displayed increased expression with time on ART for both PrEP and PWH groups (**Fig. 4c and 4d**). Intriguingly, expression of *TP53* (p53) was decreased while *MDM2* (p53 inhibitor) expression was increased with time on ART in PWH suggesting an imbalance of DDR in AM in PWH (**Fig. 4c**). Moreover, key inducers of apoptosis such as *BBC3* (i.e. PUMA) and *BAX* had decreased expression with time on ART while apoptosis inhibitors via caspase neutralization (e.g., *BCL2*, *XIAP*, and *BIRC2*) had increased expression. Two independent terms captured the SASP state of AM including the SenMayo, a well-established senescence gene set across tissues and species (**Fig. 4c**). In SASP high levels of interleukins (IL-1 and IL-6 family), chemokines (CCLs and CXCLs), and MMPs are a sign of accelerated cellular aging. Multiple SASP cytokine genes had increased expression with time on ART in AM from both the PrEP and PWH group (**Fig. 4c**). The effect of time on ART on SASP key genes such as *CXCL8, IL6R* and *IL36B* (IL-1 family member) were among the most significant changes at the transcriptomic and epigenetic levels (**Fig. 4d; Table S4**). Taken together, our analysis of the effect of time on ART found that AM of both groups displayed the epigenetic and transcriptomic characteristic of cells undergoing cell cycle arrest senescence, reduced apoptosis with dysregulated DDR and a pro-inflammatory SASP profile.

When studying time on therapy as a variable there is potential for collinearity with chronological age which may not be fully accounted for in statistical modeling. To address this, we evaluated the correlation between time on ART and the chronological age of participants. No significant correlation was detected between the participants’ chronological age and time on ART in PrEP (*R* = 0.001, *p* = 1) with a trend detected for PWH (*R* = 0.40, *p* = 0.11) (**Fig. S5a**). Next, we assessed if the chronological age was a confounder in the association of senescence and TNF signaling pathways with time on ART in PrEP and in PWH. We compared individual GESECA scores of these two pathways with chronological age in AM from both groups and failed to detect any significant correlation (**Fig. S5b**). Another possible confounder for the inflammatory and cell cycle arrest senescence-like status of AM is smoking. When comparing individual GESECA scores for senescence and TNF signaling pathways we found no association with smoking status (**Fig. S5c**). These observations reinforced the accuracy of our modeling in accounting for confounders related to pathways associated with ART. Yet, this does not exclude the independent contribution of ageing and smoking on the physiological state of AM, which was not evaluated in the context of this study.

### Increased chromatin accessibility in AM with continuous exposure to ART is enriched for motifs and footprints of pro-inflammatory and senescence associated TFs

Given that the effect of time on ART was detected in both the PrEP and PWH groups for multiple pathways, we tested the overall impact of continuous exposure to ART. We treated the HC group as a time zero and assessed the effect of time on ART on AM physiology by combining all samples while adjusting on the HC, PrEP and PWH status. We detected significant changes at FDR < 10% with continuous exposure to ART in 26.7% of the regions with accessible chromatin, 22.4% of H3K27ac, 21.8% of H3K4me3, and 52.7% of expressed genes (**Fig. 5a; Table S3**).We then repeated the GESECA analysis for the timepoints irrespective of the groups to recapitulate the correlation of biological pathways with time on ART (**Fig. S6a**). For example, the TNF signaling pathway correlated with the continuous exposure to ART, recapitulating the per group time effect (**Fig. S6b**). To showcase the effect of time on ART at the epigenetic level, we derived chromatin traces representing promoter accessibility and active histone marks for two AP-1 genes (*FOSL1*, *JUN*) and two AP-1 cooperators (*ATF3*, *ETS2*) (**Fig. 5b**). These genes showed significantly increased expression with time on ART in AM from both PrEP and PWH (**Fig. 4c)** and consequently in the combined analysis for continuous exposure to ART (**Table S3**).

**Fig. 5.**
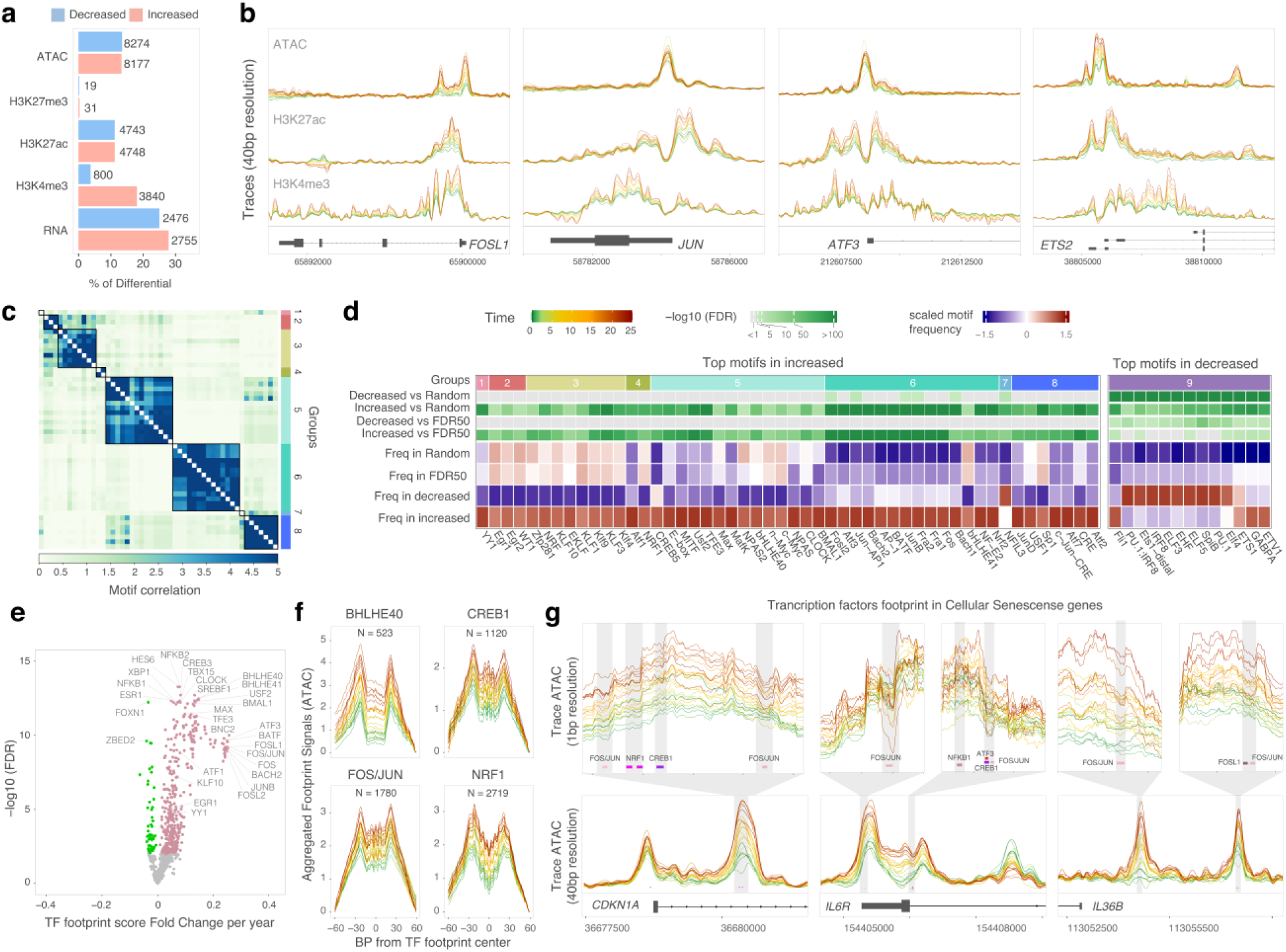
The effect of continuous exposure to ART on the epigenetic regulation of AM physiology. (**a**) Regions and genes affected by continuous exposure to ART. The bar plots show the proportion of affected regions/genes at an FDR < 10% relative to the total number of tested regions/genes. The absolute number of affected regions/genes are shown next to the bars. (**b**) Traces for chromatin accessibility, H3K27ac, and H3K4me3 marks at the promoter of four transcription factors (TF) of the TNF signaling pathway. Each line represents the normalized and covariate-corrected coverage for timepoints at a 40 base-pair (bp) resolution. (**c**) Hierarchical clustering the top 50 TF motifs found in regions with increased accessibility. (**d**) Motif frequency of the top TF motifs in the Decreased and Increased chromatin accessibility regions with continuous exposure to ART and the two control groups. The first row orders the motif-binding homology groups from “c”. (**e**) TF footprint depth in regions with increased accessibility with continuous exposure to ART. (**f**) Examples of aggregated footprint signals for TFs associated with continuous exposure to ART. Each line represents a timepoint of ART exposure. The traces depict the average signal surrounding motifs for four significant TF. The signal was normalized for the minimum signal per sample to start at 0. (**g**) Trace plots showing footprints of AP-1 subunits and cooperators in the promoter of genes associated with senescence. A dampening in the ATAC trace signal (i.e. valleys) indicates footprints on top of the motifs.

**Fig. 6.**
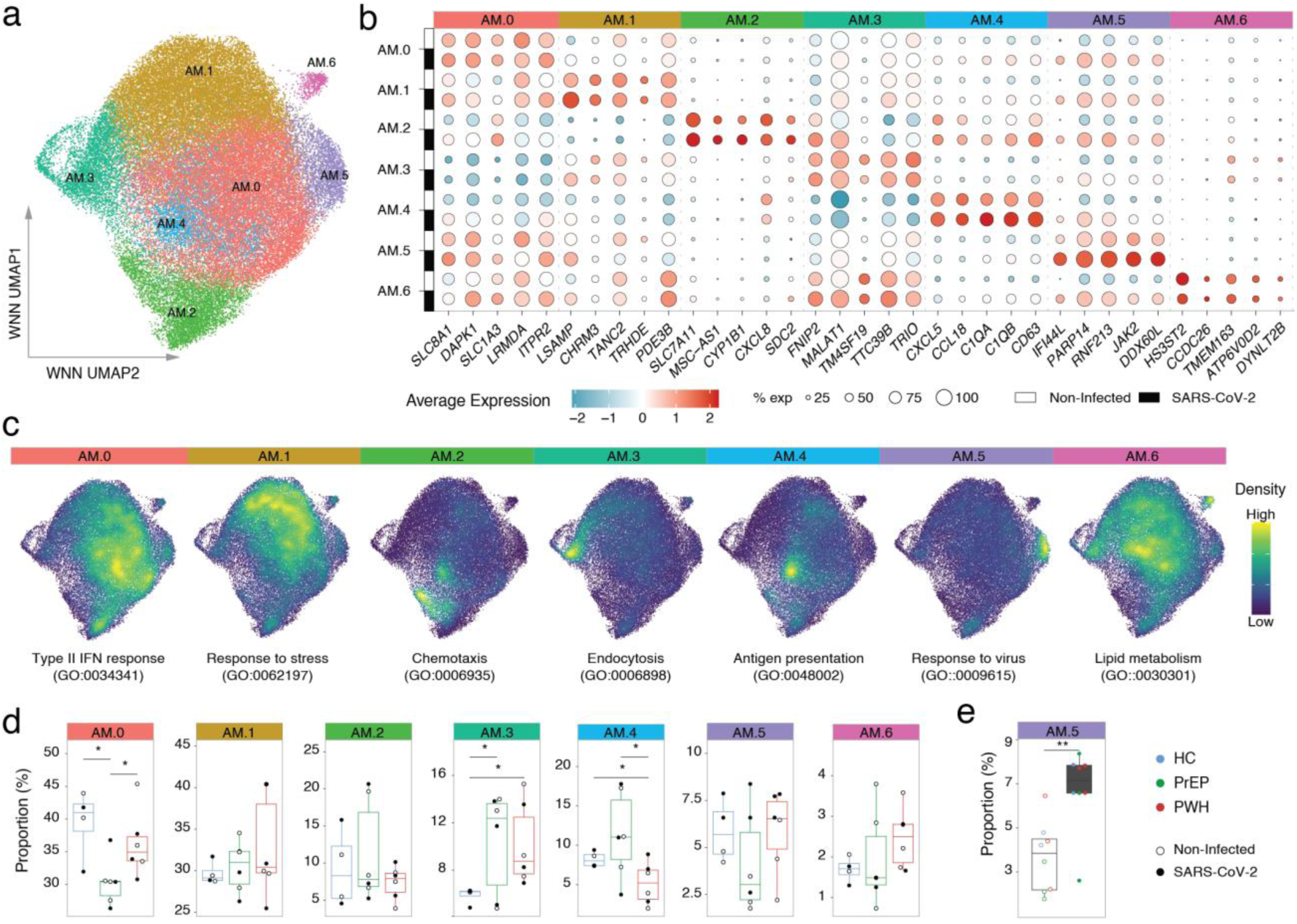
Participants on PrEP and PWH display different proportions of AM subpopulations compared to healthy controls. (**a**) snMulti Uniform Manifold Approximation and Projection (UMAP) of 60,118 cells from HC, PrEP and PWH participants either untreated or challenged with SARS-CoV-2. (**b**) Top 5 cluster markers for each of the seven AM subpopulations split by SARS-CoV-2 challenge. (**c**) AM subpopulation physiological state. A module score was calculated per cell using cluster markers assigned to the top GO term of each AM subpopulation. The colours indicate the density of cells with high module scores for the given gene ontology terms. (**d**) Proportion of AM subpopulations per group. Proportions are shown for unchallenged, and SARS-CoV-2 challenged samples. (**e**) Increase in the AM5 cell proportion following SARS-CoV-2 challenge. * *p*-value < 0.05, ** *p*-value < 0.01.

The focus of the combined analysis was to evaluate the overall epigenetic regulation of ART on the AM physiological state. Since expression of transcription factor (TF) genes, including AP-1 family members, correlated with ART duration in AM from both PrEP and PWH, we tested if TF motifs and footprints were enriched in chromatin regions showing differential accessibility with continuous ART exposure.. First, we evaluated if chromatin accessibility changes with continuous exposure to ART were enriched for motifs of TFs that supported a poised pro-inflammatory and senescent state of AM. We estimated the frequency of TFs motifs cataloged by HOMER in two control groups: (i) 500,000 random genomic regions (Random), and (ii) 20,060 ATAC regions with FDR > 50% (FDR50) for the continuous exposure to ART. Next, we compared motif frequencies in the control groups with the 8177 increased and 8274 decreased differential chromatin sites with continuous exposure to ART (ATAC at **Fig. 5a and Table S4**). We found 283 and 15 motifs significantly enriched (FDR < 1%) in regions with increased and decreased accessibility with time on ART compared to the two control groups, respectively (**Table S4**).

Next, we performed TF binding clustering by motif similarities and identified 8 groups encompassing the top 50 motifs with higher frequency in regions with increased accessibility with continuous exposure to ART (**Fig. 5c**). Group 2 included motifs for the Early growth response (EGR) family; Group 3 motifs for members of the KLF family; Group 5 motifs for MYC and MAFK; Group 6 motifs for AP-1 subunits such as Fra1 (i.e *FOLS1*), FOSL2 and JUN, and AP-1 cooperative TFs such as ATF3 and BACH; and Group 8 included members of the CRE and ATF family which crosstalk with multiple members of the AP-1 complex (**Fig. 5d**). The majority of TFs included in the five largest groups displayed coordinated effects where TFs with motifs enriched in chromatin regions with increased accessibility also showed differential gene expression with time on ART (**Table S3; Fig. S7**). The enrichment of motifs in regions with decreasing accessibility with time on ART was low. We found that 13 out of the 15 motifs with higher frequency in regions with decreased chromatin accessibility versus FDR50 were part of a single group including IRF and ELF genes (**Fig. 5d**, right).

Next, we tested the differential TF occupancy (i.e. footprint) against continuous exposure to ART. Of note, participants with the exact same time on ART were treated as a single timepoint. TF footprints were detected via depletion of ATACseq signal overlapping TF motifs within higher flanking ATACseq signals (i.e. valleys). A footprint score per timepoint was calculated for 919 JASPAR core motifs over the 8177 regions with increased chromatin accessibility with continuous exposure to ART. We then tested if the footprint mean score per TF (which is the average of footprint scores in all motifs for a given TF) correlated with the continuous exposure to ART (**Table S5**). We identified a significant increase in mean footprint score per year for 421 TFs and a significant decrease for 55 TFs (FDR < 1%) (**Fig. 5e**). Among the TF footprints with the most pronounced fold change per year (FC > 0.2) were members of the AP-1 family (**Fig. 5e**). Moreover, members of all motif groups that displayed higher frequency in increased chromatin accessibility with time on ART had significantly stronger footprints with time on ART (**Fig. 5d and 5e**). Compared to changes in TF footprint, the effect of continuous exposure to ART was more apparent for chromatin accessibility (**Fig. 5f**). Yet, most striking was the coordinated combined effect of accessibility and footprints and gene expression. For example, footprints for AP-1 subunits and other TFs associated with time on ART were identified in the core promoter of important genes mediating immune and senescence phenotypes. We detected footprints for the AP-1 JUN/FOS complex and other AP-1 cooperators in the TSS of *CDKN1A* (p21), *IL6R* and *IL36B* that are key mediators of senescence (**Fig. 5g**). These footprints had a higher depth for footprint valleys in longer ART regimens (**Fig. 5g**).

### Different proportions of distinct AM physiological states in PrEP and PWH

We hypothesized that differences in the proportions of AM subpopulations of distinct physiological state contribute to the epigenetic and transcriptomic group differences and the effects associated with continuous exposure to ART. Given the time-independent effect of ART on interferon signaling, we also hypothesized that ART impaired AM response to a virus challenge, as previously observed for *Mtb* (17). To test these hypotheses, we used snMulti to identify differences in AM subpopulation between HC, PrEP, and PWH, including their *ex vivo* response to SARS-CoV-2 challenge, and the effects of continuous exposure to ART. We selected three participants per group, prioritizing those with long-term ART in the PrEP and PWH groups (**Table 1**).

Quality control for the snMulti data was performed independently for the RNA and ATAC modalities. Nuclei that passed QC in both modalities were used for multimodal integration to identify AM clusters (**Fig. S8**). Seven distinct AM subpopulations, or “physiological states,” named AM.0 to AM.6, were identified (**Fig. 6a**). These subpopulations were independent of the SARS-CoV-2 challenge, as the leading cluster markers were observed in both untreated and SARS-CoV-2–exposed AM (**Fig. 6b**). To functionally annotate the state of each AM subpopulation, we performed gene ontology (GO) enrichment analysis using cluster markers with FDR < 5% expressed in >25% of the AM subpopulation in unchallenged samples to avoid potential bias from the SARS-CoV-2 challenge. The top GO term per subpopulation defined the main physiological state of each AM cluster (**Fig. 6c**).

**Fig. 6.**
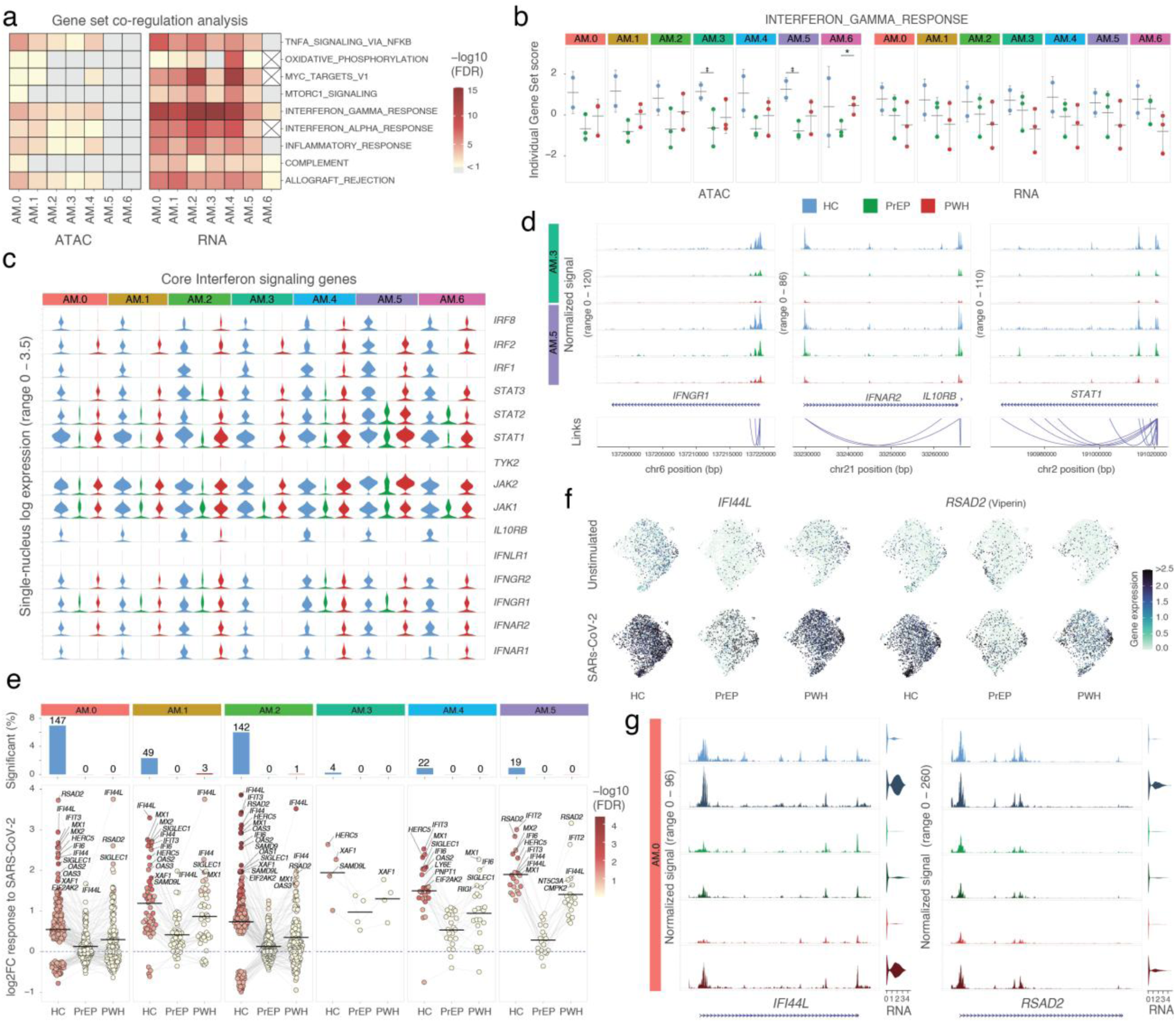
Constitutive and SARS-CoV-2–induced interferon signaling is impaired in AM from participants on ART. (**a**) GESECA hallmark gene sets with the largest variability between individuals. Gene sets with FDR < 10% in at least two assays for the same AM subpopulation are shown as a heatmap. **(b)** GESECA scores per subject and AM subpopulation separated by group for the interferon gamma response gene set. * t-test *p*-value < 0.05. (**c)** Single-nucleus expression of core interferon signaling genes. Violin plots indicate the per cell expression of the genes listed on the right. **(d)** Chromatin accessibility plots for three selected interferon response genes in the AM.3 and AM.5 subpopulations. Tracks display peaks representing baseline chromatin accessibility down sampled to show the same number of cells per group. The links indicate the correlation of peaks and gene expression. **(e)** Differential transcriptomic response to SARS-CoV-2 challenge. The top panel shows the proportion of differentially expressed genes at FDR < 10% relative to the number of tested genes for each AM subpopulation with the absolute number indicated above each bar. The bottom panel displays genes differentially expressed in at least one group for each AM subpopulation plotted across all phenotype groups by their log2 fold change in response to SARS- CoV-2. AM.6 was omitted due to absence of significant changes in any of the groups. The crossbar indicates the median fold change, including both upregulated and downregulated genes. **(f)** snMulti UMAP with gene expression of two genes of the interferon gamma gene set. **(g)** Chromatin accessibility plots for the genes shown in panel f. Tracks were generated as in panel d and separated by group and SARS-CoV-2 challenge status. Single-nucleus expression levels for each gene are shown on the right for each condition.

To assess AM state transitions and their modulation by group or SARS-CoV-2 challenge, we performed trajectory and pseudotime analyses. In both approaches, the trajectories were not altered by either phenotype group or SARS-CoV-2 challenge (**Fig. S9**). Next, we quantified the proportion of AM subpopulations across phenotype groups (**Fig. 6d**). Participants on PrEP had a significantly lower proportion of cells in the type-II interferon subpopulation AM.0 while PWH had a significantly lower proportion in antigen presentation AM.4. Both PrEP and PWH showed a significantly higher proportion of the AM.3 state which is characterized by endocytosis and is depleted of interferon responsive genes (**Fig. 6d**). Only subpopulation AM.5 significantly increased in proportion following *ex vivo* SARS-CoV-2 challenge, independent of phenotype group (**Fig. 6e**)

### Constitutive impaired interferon signaling in AM physiological states enriched in participants on ART

We confirmed the main biological pathways associated with high inter-individual variability observed in the bulk experiments in the snMulti (**Fig. 2b** and **Fig. 7a**). Using a pseudobulk approach per AM subpopulation we observed that the most variable gene sets between participants included interferon responses, MYC targets, OXPHOS, and TNF signaling via NFKB. This was consistent with the bulk-level findings. The interferon gamma response gene set was equally significant across most AM subpopulations while other gene sets were differently distributed across specific AM subsets (**Fig. 7a**). Module scores per sample for the interferon gamma gene set indicate a global trend of reduced chromatin accessibility and gene expression in participants on ART independent of infection (**Fig. 7b**). Notably, participants on PrEP showed significantly lower module scores for chromatin accessibility in AM.3 and AM.5, subpopulations representing the lowest and highest interferon response states, respectively (**Fig. 7b**). This constitutive impairment extended to core components of the interferon signaling pathway not included in the hallmark gene set. For example, the single-nucleus expression of key mediators of type I, II, and III interferon signaling has constitutively lower nuclear expression in AM from PrEP, and to a lower extent from PWH, compared to HC (**Fig. 7c and Table S6**). This finding is in line with the time-independent reduction of *IFNGR1* and *IFNAR2* expression detected in the cytoplasm of AM from PrEP at the bulk stage (**Fig. 2e**). The transcriptional repression of interferon signaling was coordinated with suppression of chromatin accessibility at the promoters of interferon receptors (e.g., *IFNGR1, IFNAR2, IL10RB*) and downstream effectors (e.g., *STAT1*), across all subpopulations, including the polar extremes AM.3 and AM.5 (**Fig. 7d**). The interferon gamma response gene set also included multiple antigen presentation genes. We observed that participants on ART had reduced nuclear expression and promoter accessibility of HLA genes (**Fig. S10 and Table S6**). This effect in HLA-DRA and the non-classical class II molecules HLA-DM was consistent across both bulk and single nucleus profile (**Table S1** and **Table S6**). Together, these results expand on the presence of a constitutively impaired interferon response state in AMs of individuals on ART that was more pronounced in PrEP and in the AM.3 physiological state, which is more frequent in AM isolated from PrEP and PWH participants. (**Fig. 6e**).

**Fig. 7.**
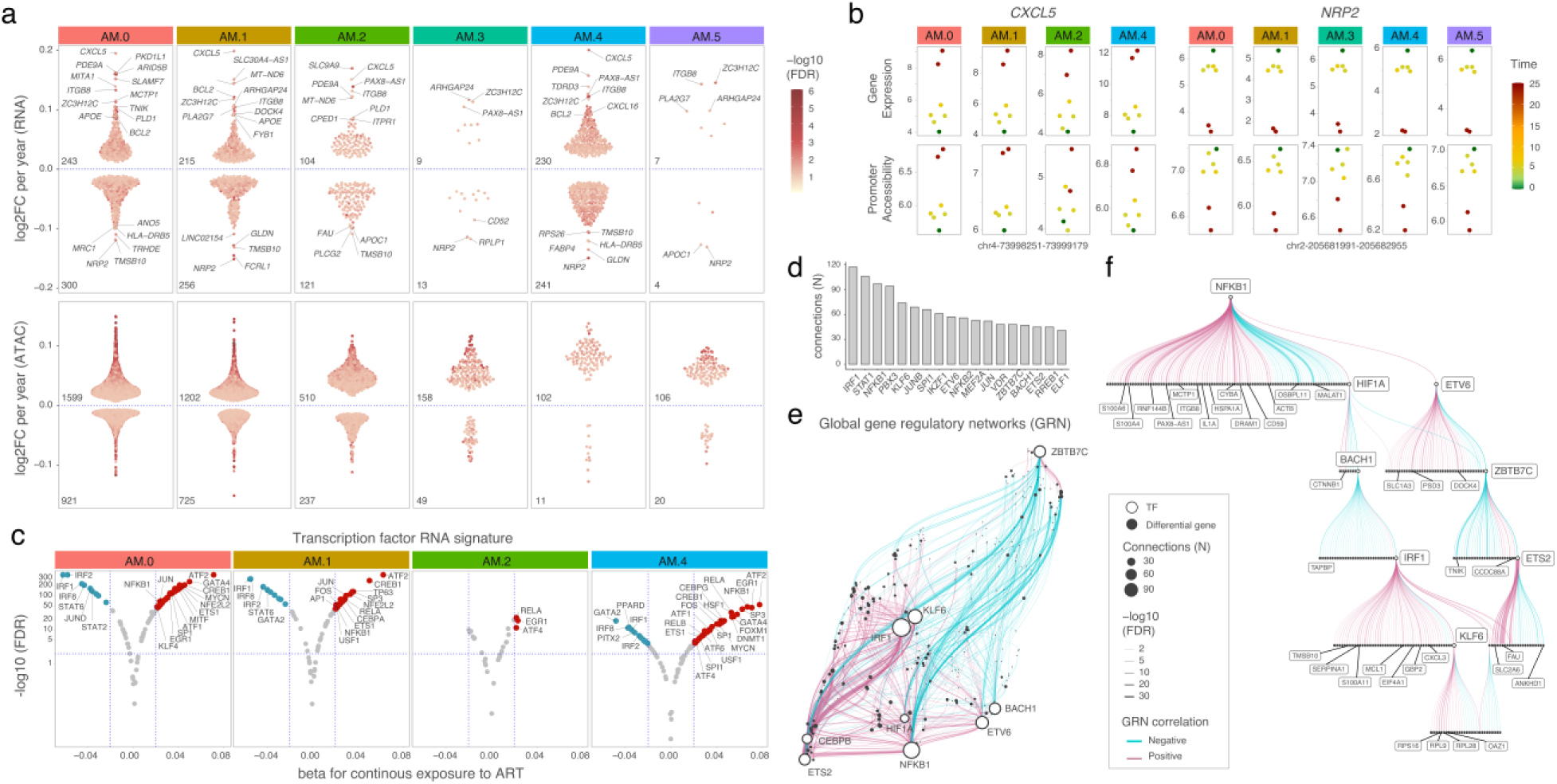
Coordinated TF activity indicates a senescence-like pro-inflammatory profile in AMs with continuous exposure to ART. (**a**) Differential features associated with continuous exposure to ART for snRNA and snATAC. Differential features with FDR < 10% were plotted according to the log2 fold change per year of ART exposure per AM subpopulations. (**b**) Top genes differentially up-(*CXCL5*) or down-regulated (*NRP2*) with continuous exposure to ART. (**c**) TF activity scores with continuous exposure to ART. The x-axis shows beta coefficients for the association between single-nucleus TF activity and continuous exposure to ART. The y-axis shows the corresponding FDR *p*-values. Red and blue indicating increased and decreased activity, respectively. **(d–f)** snMulti Gene Regulatory Network (GRN) analysis based on the 3,332 unique differential chromatin regions and 867 differential genes identified in panel “a”. (**d**) The y-axis indicates the number of differential genes (N) connected to differential regions containing motifs for the corresponding TFs shown on the x-axis. Only TFs with more than 50 significant connections are displayed. (**e**) Global GRN module. The network shows connections between TFs and their associated target genes. Edge color indicates the direction of the correlation and edge width the statistical FDR of the connection. (**f**) TF subnetwork highlighting NFKB1 position at the top of a cascade within the GRN. Each circle in the hierarchical tree represents a target gene of the TF network.

### Impaired AM response to SARS-CoV-2 ex vivo challenge in participants on ART

Interferon signaling is a critical component of host defense against viral pathogens. Hence, we tested if the constitutively reduced interferon signaling observed in AM in participants on ART affected their ability to mount a robust epigenetic and transcriptomic response following *ex vivo* SARS-CoV-2 exposure. AM from HC showed significant transcriptional changes in response to the virus while both PrEP and PWH showed few or no significant changes passing FDR < 10% (**Fig. 6e**, top and **Table S7**). Across all AM subpopulations, the magnitude of nuclear transcriptional response to SARS-CoV-2 was remarkably lower in participants on ART for the differentially expressed genes (**Fig. 6e**, bottom). As expected, interferon related genes displayed the highest fold changes in response to SARS-CoV-2. The most strongly induced genes were *IFI44L* and *RSAD2* (**Fig. 6e** and **6f**). In contrast, we detected only a few significant chromatin accessibility changes following SARS-CoV-2 exposure, none of which were enriched in a specific gene set. This lack of epigenetic response may reflect both the small sample size, number of tests and the sparsity of snATAC data when analyzed at the pseudobulk level. Nevertheless, we observed changes in chromatin accessibility at the promoters of interferon response genes, including *IFI44L* and *RSAD2* (**Fig. 6g**). However, baseline differences in promoter accessibility between groups were substantially more pronounced than the changes induced by SARS-CoV-2 (**Fig. 6g**).

### Single nucleus TF signature of senescence-like pro-inflammatory profile with continuous exposure to ART

We investigated if the senescence-like pro-inflammatory profile associated with continuous exposure to ART was restricted to specific AM subpopulations. To address this, we performed pseudobulk differential feature analysis for continuous exposure to ART stratified by AM subpopulation. At the nuclear transcriptome level, subpopulations AM.0, AM.1 and, AM.2 had the highest proportions of differentially expressed genes with continuous exposure to ART (**Fig. 7a**, top, and **Table S8**). Interestingly, AM.3 displayed fewer differentially expressed genes compared to AM.4. Considering that AM.3 encompass more cells than AM.4 it confirms the IFN attenuated physiological state of the subpopulation enriched in PrEP and PWH in response to SARS-CoV2. Among the top differentially expressed genes, the pro-inflammatory chemokine *CXCL5* showed the highest fold change per year on ART, while the immunomodulatory *NRP2* showed the lowest (**Fig. 7b**). The expression of both genes was accompanied by corresponding changes in promoter accessibility (**Fig. 7b**). In contrast, the count of differentially accessible chromatin decreased accordingly to the number of cells per subpopulation (**Fig. 7a**, bottom, and **Table S8**). Several genes and chromatin regions associated with continuous exposure to ART at the bulk stage were below detectability and could not be tested. Nevertheless, at the AM subpopulation level, we observed the same trend of association with continuous exposure to ART for key senescence markers, including *CXCL8* and the IL6 transducer *IL6ST* from SASP, as well as *MDM2*, *BCL2,* and *E2F3,* which are involved in distinct mechanisms of senescence (**Fig. S11**).

We investigated the extent to which the senescence-like pro-inflammatory TF signature associated with continuous exposure to ART at the bulk stage was captured by a specific AM subpopulation. For that, we inferred TF activity based on the expression of known target genes either induced or repressed by each TF. Using the differentially expressed genes identified per subpopulation shown in Fig. 7a, we calculated TF activity scores per nucleus and tested their association with continuous exposure to ART per AM subpopulation using a linear model. TF with absolute beta > 0.02 and FDR < 1% were considered significant (**Fig. 7c**). The results provided confirmation at the single nucleus level of the bulk findings and indicate that the continuous exposure to ART effect is not restricted to one AM subpopulation. We observed increased TF activity for AP-1 family members (e.g., JUN and FOS) as well as other TFs identified through differential footprint analysis in the bulk stage (e.g., NFKB1, CREB1, ATF2, EGR1) with continuous exposure to ART (**Fig. 7c**). Conversely, interferon-responsive factors (IRFs) showed decreased activity, consistent with the reduced motif enrichment observed with continuous exposure to ART in the bulk stage (**Fig. 4d** and **Fig. 7c**). The senescence-like TF signature was more pronounced in subpopulations AM.0, AM.1, and AM.4. However, this may be a reflection of the higher number of differentially expressed genes which are used to assess TF activity.

The TF activity analysis considered only the snRNA modality. To harness the full power of snMulti we performed a TF gene regulatory network (GRN) analysis using the differential features from the ATAC and RNA modalities shown in Fig. 7a. For each TF we modeled gene expression as a function of the interaction between chromatin accessibility at regions containing the TF motif and the TF’s expression across single nuclei using Pando framework. TF–target gene connections with FDR < 10% were considered significant. We performed the analysis for all AM subpopulations combined. TFs with more than 50 significant connections included the core regulator of pro-inflammatory signaling (NFKB1) and senescence-associated TFs (e.g., JUN, ETS2 and KLF6) (**Fig. 7d)**. Next, we estimated a global GRN module comprising co-regulated TF–target gene signatures (**Fig. 7e**). This network reflects the coordinated effect of key TFs in regulating differential gene expression. A central TF in the global GRN was NFKB1, which, together with HIF1A, ETS2, and KLF6, underlines the senescence-like pro-inflammatory state of AM at the single-nucleus level (**Fig. 7f**). Establishment of the TF hierarchy allows the targeting of key TF for improved or adjuvant therapy.

A limitation of the snMulti GRN approach is that it strongly relies on TF expression modeling, which can be challenging in highly differentiated cells like AM with low baseline TF expression, and number of nuclei. As a result, GRN analysis stratified by subpopulation was feasible only for larger subpopulations such as AM.0 with sufficient number of nuclei. Nevertheless, in AM.0 we captured part of the GRN modules identified in the combined AM subpopulations. Importantly, the senescence-like pro-inflammatory signature driven by NFKB1, ETS2, and KLF6 was still observed, likely because it was not specific but rather shared across multiple AM subpopulations. This suggested that the senescence-like pro-inflammatory profile was not restricted to a single AM subpopulation and was consistent with both the bulk-level observations and the TF activity inferred from snRNA. Taken together, these results reinforced the presence of a TF signature consistent with a senescence-like pro-inflammatory profile in AMs of participants with continuous exposure to ART.

## Discussion

We found two major effects of NRTI-based ART in AM. First, interferon responses were impaired both at basal and following a viral challenge with stronger effect in HIV-negative PrEP participants and independent of ART duration. Second, continuous ART exposure reshaped AM homeostasis and cellular state at the epigenetic and transcriptomic levels. Both PWH and PrEP participants showed a progressive senescence-like pro-inflammatory profile that correlated with duration of NRTI exposure.

Cellular senescence encompasses multiple biological processes and is often referred to as the “zombie cell” state, in which cells are under growth-arrest and resistant to apoptosis while remaining metabolically and transcriptionally active. These cells are viable despite their cytotoxic microenvironment but display an impaired ability to execute their proper physiological role. In our study, AM from both PrEP and PWH participants exhibited epigenetic and transcriptomic features consistent with this state. Genes involved in cycle arrest, such as *CDKN1A* (p21), *CHEK2*, and *GADD45A* had increased expression with longer ART duration, while *GADD45B* expression was elevated in both PrEP and PWH AM, independent of time on ART. These proteins mediate G2 phase arrest, preventing progression to the M phase (52–54). Concurrently, key pro-apoptotic inducers such as *BAX* and *BBC3* (PUMA) were downregulated, while anti-apoptotic genes such as *BCL2*, *XIAP* and *BIRC2* were upregulated with increased time on ART. Together, these gene expression changes suggest that viable AM are at least partially trapped in the G2 phase which combined with the cellular and mitochondrial stress and pro-inflammatory SASP profile could be detrimental to their cellular function. Another aspect is paracrine senescence in which SASP cytokines propagate the senescent state to neighboring cells (55). In this context, the effects of continuous NRTI-based ART are likely not exclusive to AM but rather systemic. Supporting this, studies in PWH on long-term ART have shown elevated plasma levels of SASP cytokines such as IL-8, TNF, MMP-1, and CCLs which is consistent with our transcriptomic findings in AM (56, 57). Additionally, NRTI exposure in astrocytes has been shown to induce p21, secretion of IL-1 family members, and activation of NF-κB p65, accompanied by increased ROS production and mitochondrial dysfunction (58). Such systemic ART effects may be undetectable during short-term ART or masked by the intertwined relationship with HIV infection. However, in analogy to the time dependency of transcriptomic changes they might accumulate over the years potentially affecting cellular function in multiple tissues. Such a detrimental effect on AM function has been shown directly through the abrogated AM responses to both SARS-CoV2 and *Mycobacterium tuberculosis* (17).

Aging PWH on ART have a higher incidence of chronic airway diseases, including COPD, lung fibrosis, and asthma, as well as a higher burden and early onset of lung cancers compared to ART-naive individuals (59, 60). AM genes displaying differential expression with time on ART in our study have been implicated in the pathogenesis of these conditions. For example, increased expression of AP-1 subunits in macrophages promotes tissue remodeling in COPD and lung fibrosis (61, 62). Moreover, SASP cytokines such as IL-6 and IL-8 were observed to be elevated in the serum and BAL fluid of patients with COPD and asthma (63–65). SASP is also characterized by increased oxidative stress and overproduction of ROS by mitochondria or reduced antioxidant capacity (59, 66). In our study, we detected an impaired expression of major antioxidant enzymes with longer time on ART. Oxidative stress-based therapeutics are under investigation as candidates to treat the underlying pathogenic mechanisms of COPD (67). Although AM are not a typically metastatic cells, they shape the tumor microenvironment in the lung (68). Increased expression of AP-1 subunits *FOSL1* and *JUN* regulated acquired resistance to treatment in non-small cell lung carcinoma (NSCLC), which was reversed with AP-1 inhibitors (69). Another key oncogenic pathway is the p53-MDM2 loop. We observed *TP53* downregulated with increased time on ART in PWH with a corresponding increased expression of its antagonist *MDM2* in both PWH and PrEP participants. *MDM2* overexpression is a common feature in lung cancers and the MDM2-p53 loop contributes to chemotherapeutic resistance in human malignancies (70, 71). More broadly, we detected impairment of DDR mechanisms and cell cycle checkpoints in AM from both PWH and PrEP, suggesting that prolonged ART may in part explain the increased susceptibility to lung cancer in PWH.

Tuberculosis, pneumonia and more recently COVID-19 are major causes of death in PWH under ART (14–16, 72). We previously showed that AM obtained from PWH and people on PrEP had a diminished transcriptomic and epigenetic response to *Mtb* and here we extended these findings to SARS-CoV-2 (17, 73–75). Interferons are key modulators of pulmonary host defense and the overall decreased IFN mediated response we observed impaired the capacity of AM to mount a robust response to these two distinct pulmonary pathogens (76). IFN neutralizing autoantibodies have been shown more frequently in people with severe COVID-19 and in life-threatening COVID-19 pneumonia, which strengthens the importance of a balanced interferon responses in the outcome of SARS-CoV-2 infection (77, 78). Of relevance is the lower IFN receptors expression in AM from PrEP participants independent of time on ART which was accompanied by an increased proportion of unresponsive AM subpopulations. While we do not know if the blunted AM response to *ex vivo* SARS-CoV-2 challenge does have a major impact on COVID-19 outcomes in people on PrEP, our results support the need for more detailed longitudinal epidemiological data to assess the clinical implications of our findings. However, assuming that an increased and wider scale of PrEP uptake may involve medicating millions of healthy people with NRTI, potential side effects need to be further pondered either through the use of alternative ART classes or with adjuvant therapies particularly in low HIV endemic settings (79).

PrEP is a highly effective strategy to prevent HIV acquisition (80). Our study does not advocate against PrEP rollout in any way as protection from HIV infection dramatically outweighs potential ART side effects. PrEP rollout in high exposure settings contributes to the decline of HIV diagnosis at the population level and discontinuation, intermittent use or low PrEP compliance are known to reduce prevention effectiveness (81–85). A few studies have highlighted adverse effects by short term exposure to the most common NRTIs tenofovir disoproxil fumarate (TDF) and emtricitabine (FTC). Short term TDF/FTC exposure induced tissue-specific IFN transcriptional signatures with upregulation of IFN-response in the gut and downregulation in PBMCs and blood (86). Leukocytes exposed to TDF/FTC displayed increased ROS production and lipid uptake and decreased mitochondrial mass and efferocytosis (87, 88). NRTI exposure in monocyte-derived macrophages caused mitochondrial dysfunction, higher levels of ROS and induction of SASP cytokines IL-6, IL-1β and TNF (89). Moreover, TDF was shown to inhibit *in vitro* telomerase activity in PBMC from PWH (90). Combined these observations on short term exposure to ART suggests a senescence-like profile in immune cells induced by NRTI. However, the mechanisms by which long-term NRTI-based ART impacts cell physiology independent of HIV remains largely unknown.

A strength of the snMulti is the ability to evaluate regulatory networks linking chromatin accessibility and transcription factors with their target genes at each individual nucleus. We applied this powerful approach to investigate the gene regulatory network that underlie the adverse effects of long-term ART. We identified key transcriptions factors in central regulatory hubs modulating the transcriptional changes associated with ART. However, targeting these key transcription factors is challenging as many hubs lack pharmacological modulators or are considered undruggable due to their essential and widespread functions such as the central hub of NFKB. However, engineered exosome delivery technologies are being developed to specifically modulate NFKB activity by delivering long-lasting super-repressor inhibitor of NFKB (Exo-srIκB). The Exo-srIκB system has been tested in rodent models of kidney ischemia (91), sepsis (92)and age-related neuroinflammation (93). While these animal studies are promising the exosome system currently lacks cell specificity. Moreover, in the context of ART systemic inhibition of NFKB may itself could cause adverse effects that outweigh potential benefits. By contrast, a possible exception among the key hubs we identified is HIF1A, for which a range of pharmacological modulators have been developed and evaluated in pre-clinical and clinical studies (94). Currently, the main clinical trials focus on ischemic disease and specific cancers rather than pathways of immune activation (95, 96). Nevertheless, targeting of HIF1A may represent an alternative or complementary approach to targeting senescence beyond senolytics in person on long term ART. While HIF1A as modifiable target is attractive, the fact that adverse ART effects in AM require prolonged exposure means that clinical trials would face extensive logistical challenges. This further supports the rationale to study ART-adverse effect in cells other than AM. In such cells adverse effects may emerge more rapidly and might provide useful biomarkers for interventional studies.

Our study has some limitations. AM are not an optimal cell type to evaluate the systemic effects of NRTI-based ART. Our rationale for focusing on AM was based on our previous observation of a blunted transcriptomic and epigenetic response to *Mtb* in both PrEP and PWH. The extent to which this localized effect in the lung reflects systemic immune alterations remains unknown. Future systems-level studies assessing circulating or progenitor cells combined with immediate clinically relevant assays (e.g., protein production) will be required to evaluate the long-term effects of ART on clinical outcomes. Moreover, of the participant in the PrEP and PWH groups only one was female. Therefore, with our dataset it is not possible to test a sex bias of the ART effect. Sex is associated with different levels of drug metabolism, which might impact the ART effect in females. Participants of the PrEP group were more homogeneous using the same NRTI ART regimen while in the PWH group, participants had more complex therapies. PWH were exposed to multiple and different types of ART drugs throughout their life. Exposure to older and more toxic ART might have contributed to the DNA damage observed in AM from PWH. Importantly, our study did not allow to address the mechanisms and the ART drug concentration that underlie the *in vivo* senescence-like pro-inflammatory state of AM.

Our study has several unique strengths. First, it is the only study to date that evaluated the long-term epigenetic effect of ART in the HIV^+^ and HIV^-^ context. Second, studies of PrEP side effects in healthy subjects have relative short follow-up periods ranging from a few months to about a year. In contrast, our retrospective cross-sectional approach evaluated a much longer effect of ART which would be challenging to implement in a longitudinal design. Additionally, participants of the PrEP and PWH groups did not include opioid users, and both groups have similar numbers of cannabis users.. Another aspect is that studies on ART focused on PWH, making it difficult to separate the HIV-effect from the ART-effect. Our study addressed this gap by studying ART effects in both HIV^+^ and HIV^-^ persons allowing us to identify specific ART-effects. Our study included three PHW with aprox. 10 years gap between HIV diagnosis an ART initiation. These participants showed an AM profile that was in line with the time on ART and not with the time from HIV diagnosis, which reinforced the role of NRTI-based ART in the AM physiological state.

Most importantly, our study highlights the constant need for improved drug designs and multidrug therapies to mitigate side effects. Treatment and prevention of HIV infection have come a long way since the introduction of the first generation of highly toxic ART (e.g., zidovudine, stavudine, didanosine). However, our study indicates the gaps that still need to be addressed towards improved treatment to ensure the best quality of life for ART users. If the discovered effect is exclusive to the NRTI class adjuvant therapies could be beneficial. For example, senolytics have been shown to mitigate cellular senescence with compounds such as dasatinib, quercetin, and fisetin currently being tested in clinical trials for pulmonary diseases linked to senescence (97, 98). Alternatively, drugs with distinct mechanisms of action could potentially circumvent the side effects of long-term NRTI-based ART. Long-lasting injectables, such as the HIV integrase inhibitor Cabotegravir and the capsid inhibitor lenacapavir, have been investigated or approved as potential PrEP regimens (99–102). Albeit for most people at risk of HIV acquisition the efficacy of PrEP outweighs the risks of potential side effects; our results suggest that lengthy and continuous exposure to ART requires a highly judicious use, particularly for diseases where AM play a critical role.

## Data Availability

The raw sequencing and processed datasets for the bulk and snMulti assays were deposited to GEO with accession numbers XXXX. Additional data are available from the corresponding author on reasonable request.

## Declarations

### Ethics approval and consent to participate

The study was approved by the Research Ethics Board of the McGill University Health Center (MUHC) (MP-CUSM-15-406). All participants signed a written informed consent and were recruited at the McGill University Health Centre (MUHC, Montreal, Canada).

### Consent for publication

Not applicable

## Acknowledgements

We thank all volunteers who participated in this study. We thank the members of bronchoscopy team at MUHC, the members of the Routy and Costiniuk labs, and all members of the Schurr lab for their help and discussions.

## Supplementary Materials

**Fig. S1.**
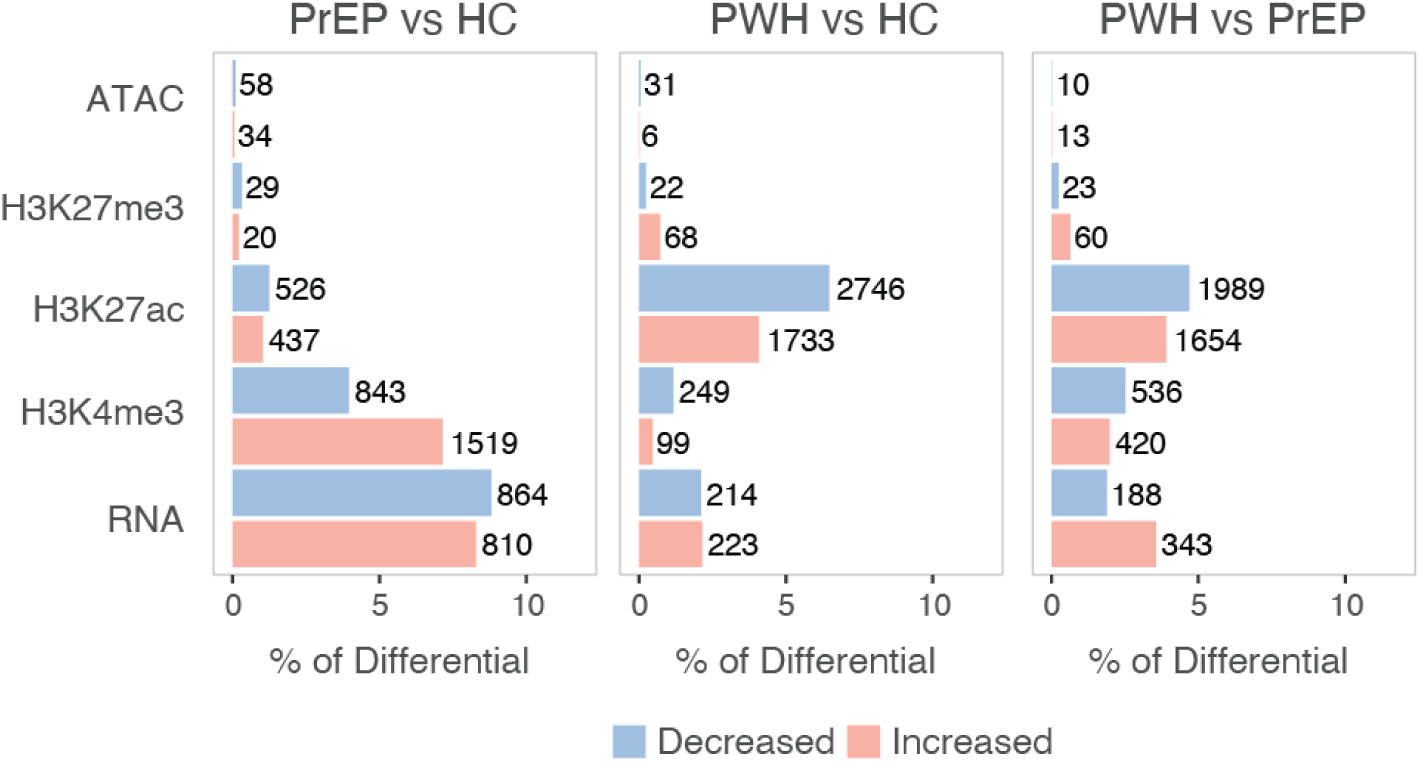
Differential epigenetic and transcriptomic features between groups. Differential epigenetic and transcriptomic features between groups are shown for the three baseline comparisons. The bar plots show the proportion of differential regions/genes at an FDR of 10% and absolute log2FC > 0.2 relative to the total number of tested regions/genes for time on ART in PrEP vs HC (left), PWH vs HC (center) and PWH vs PrEP (right). The absolute number of differential regions/genes are shown next to the bars. Decreased and increased DNA accessibility, histone marks, and gene expression are shown in blue and red, respectively.

**Fig. S2.**
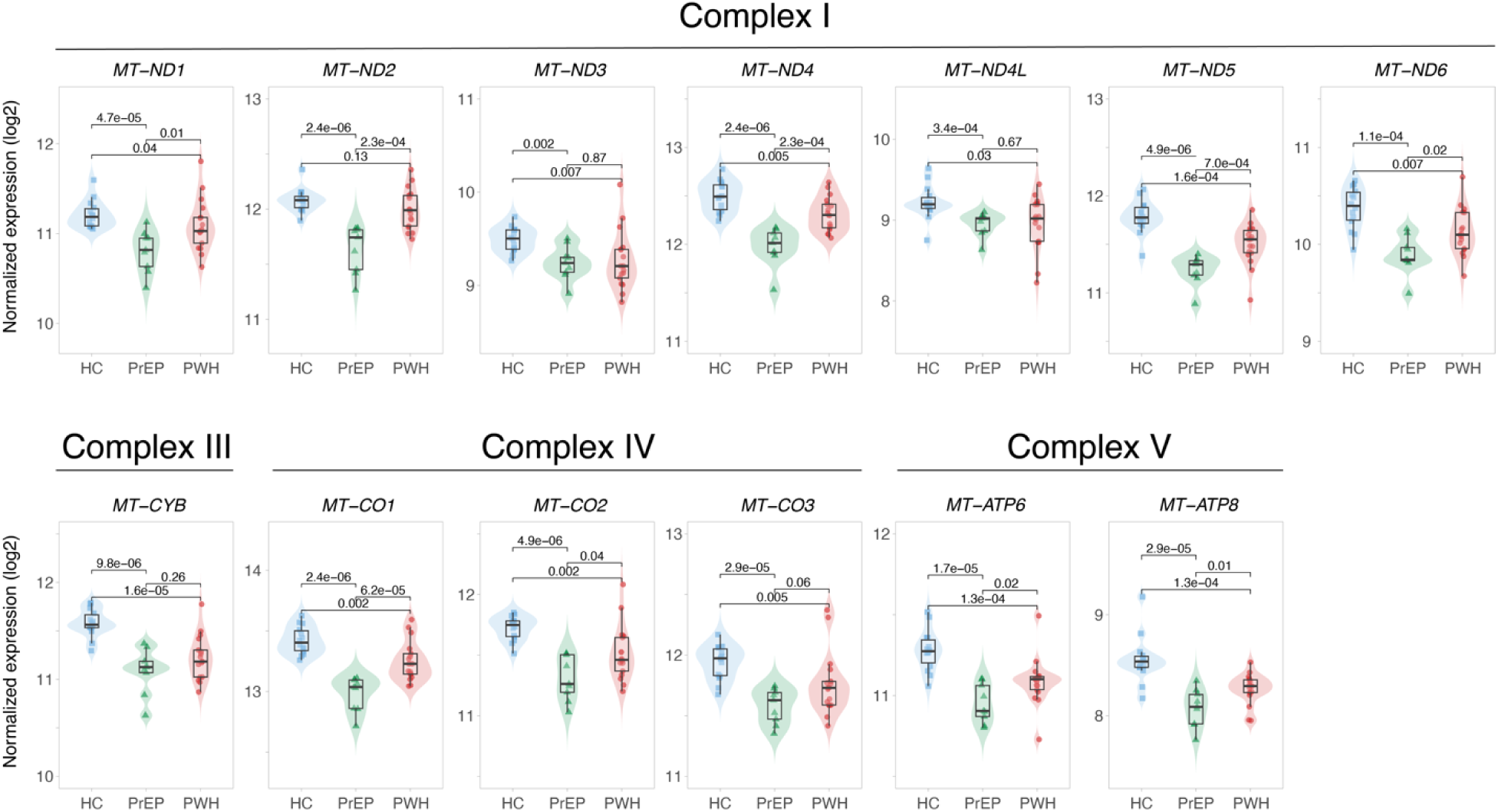
Reduced expression of mitochondrial genes encoding enzymes of the electron transport chain. Normalized gene expression per subject and group for mitochondrial genes encoding proteins of the human electron transport chain. *p*-values for the Wilcoxon test comparing differences in median expressions between groups are shown at the top.

**Fig. S3.**
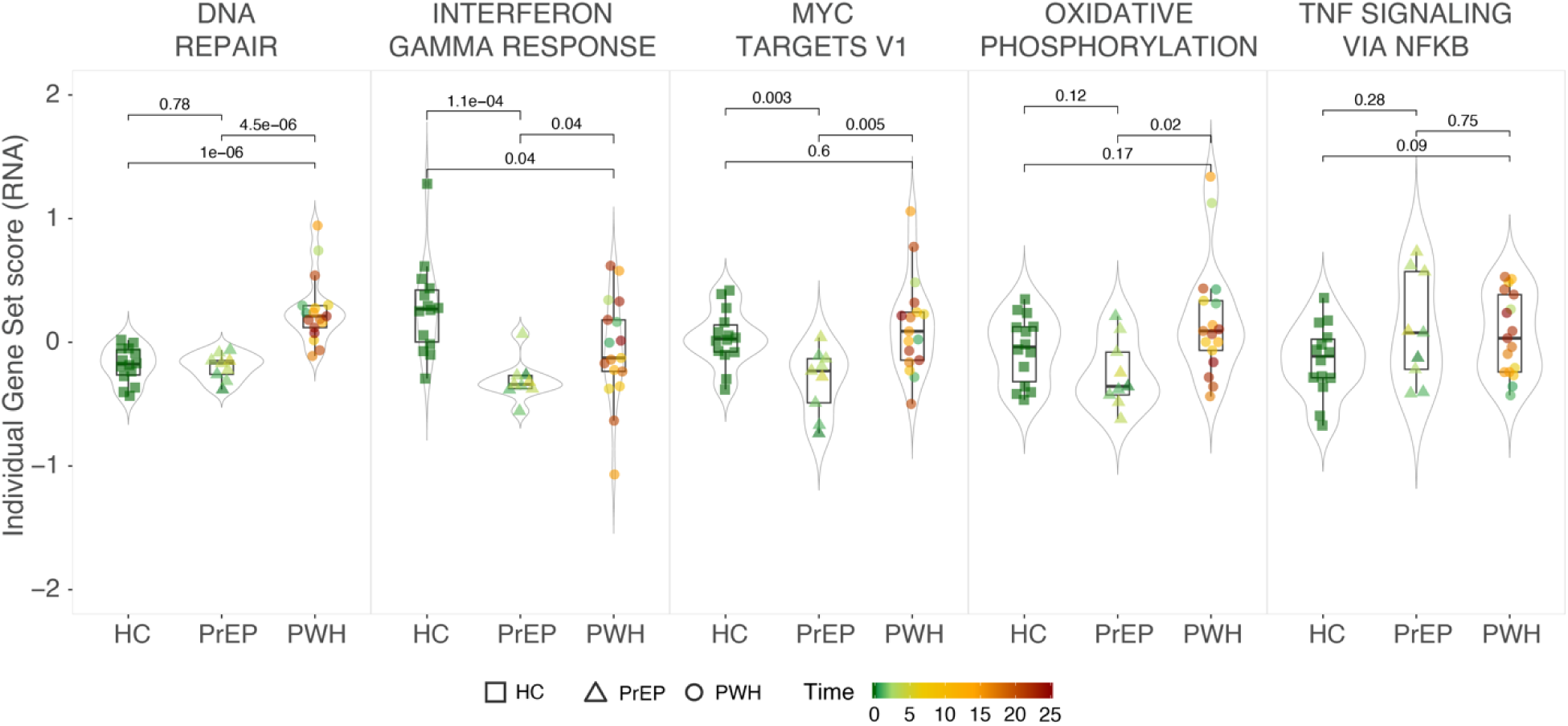
Comparison between GESECA scores and phenotypic groups as in Fig. 2C indicating time on ART. GESECA scores for the transcriptomic assay for each subject are plotted on the y-axis separated by phenotypic group on the x-axis. The *p*-values shown at the top are for the group compassions shown in Fig. 2C. The colour shades indicate the length of time on ART for PrEP and for PWH.

**Fig. S4.**
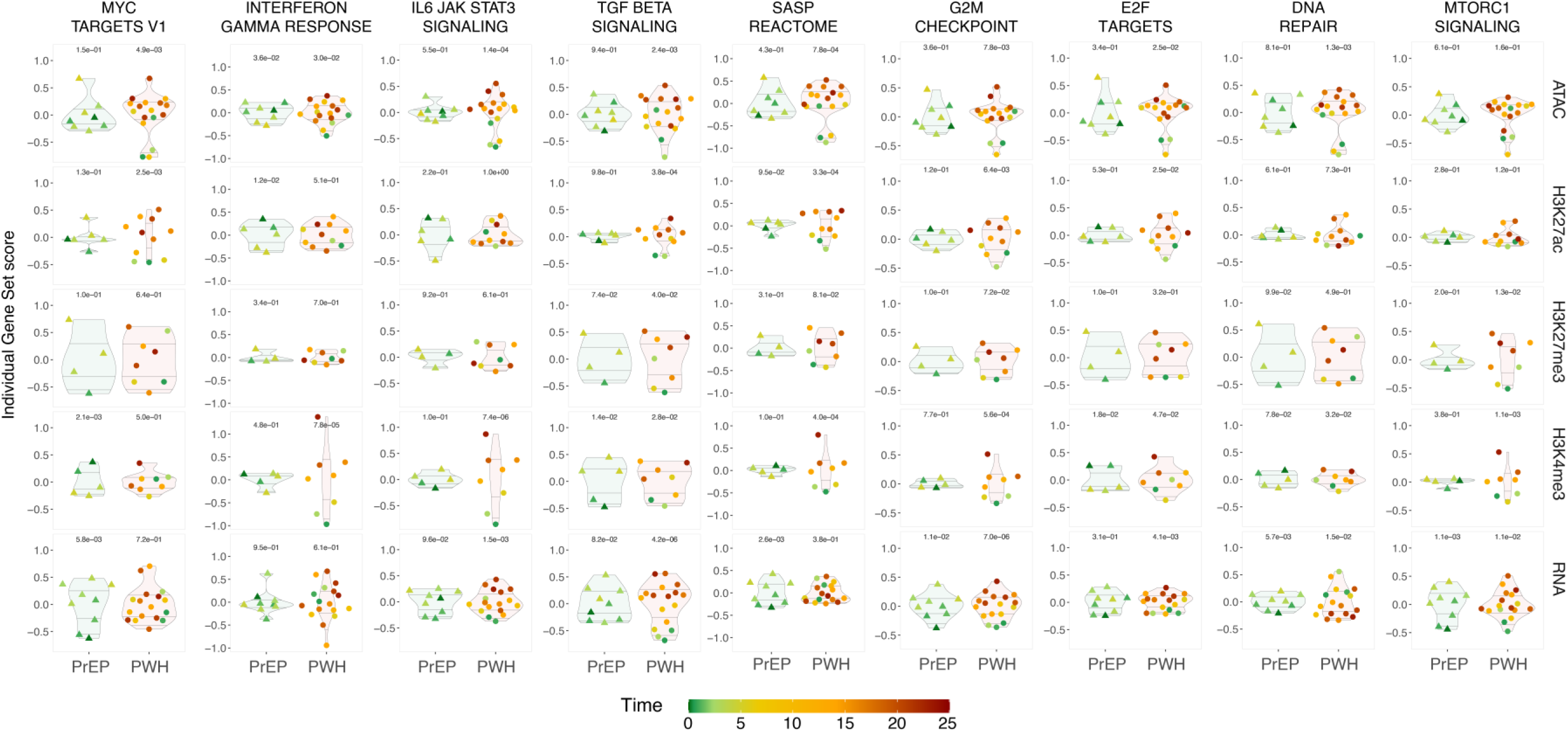
Per subject GESECA score for eight pathways affected by time as a complement to Fig. 4B. Each dot represents an individual with colour shading indicating time on ART for PrEP or for PWH. GESECA scores per individual were plotted on the y-axis and subjects were separated by groups on the x-axis. The quantiles are shown as violins for reference of the distribution. A linear model was used to test the correlation of time on ART on PrEP or on PWH with the individual GESECA scores for all tested assays. *P*-values for the correlations are shown at the top of each comparison.

**Fig. S5.**
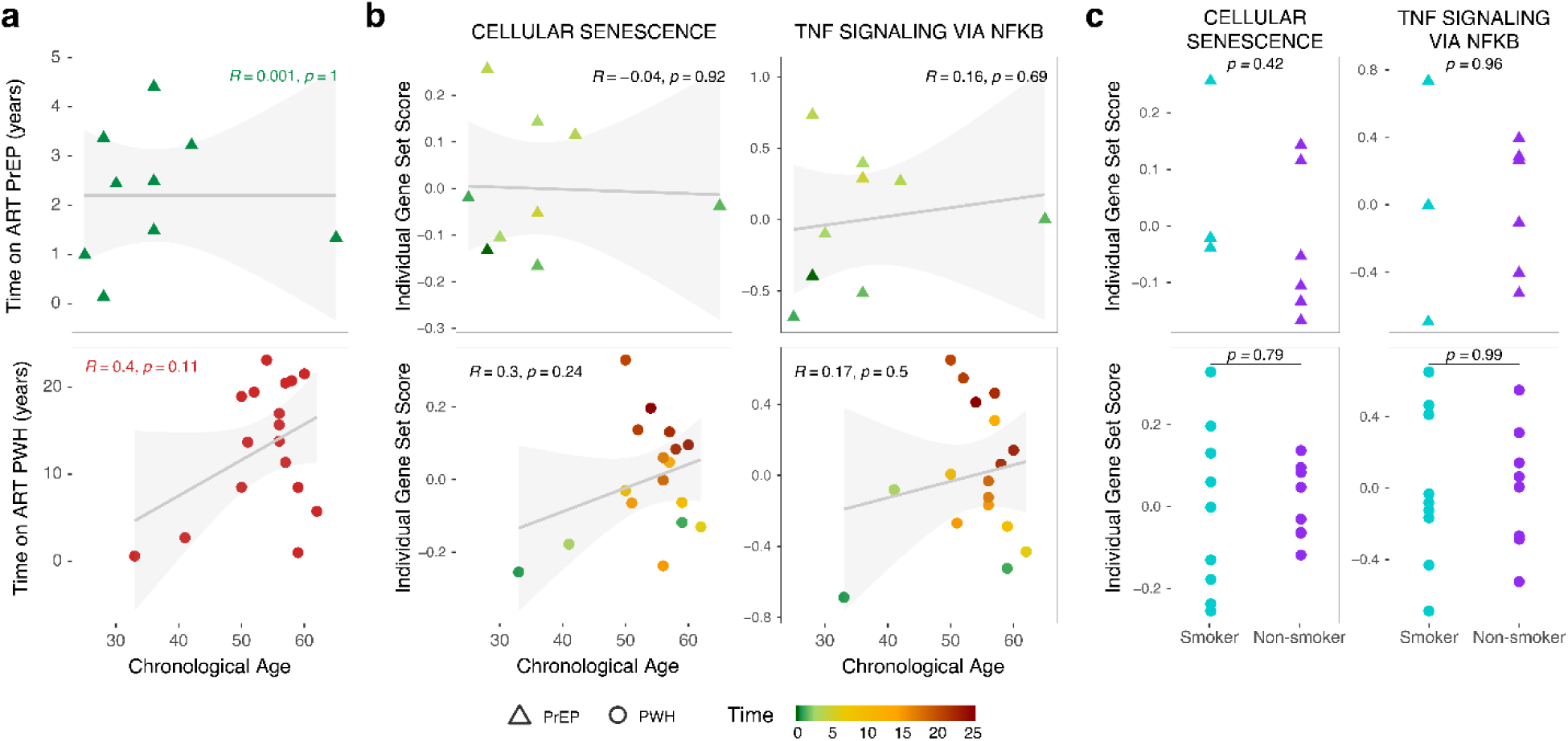
Effect of chronological age and smoking status on ART associated pathways. (**a**) Correlation plot between chronological age and time on ART for people on PrEP (top) or for PWH (bottom). (**b**) Correlation plot between individual gene set scores of the RNA assay for two main pathways associated with time and chronological age. (**c**) Comparison of individual gene set scores of the RNA assay and smoking status for two main pathways associated with time on ART. PrEP subjects are shown on top and PWH at the bottom graphs. The *p*-values indicate differences in the mean GESECA scores between groups.

**Fig. S6.**
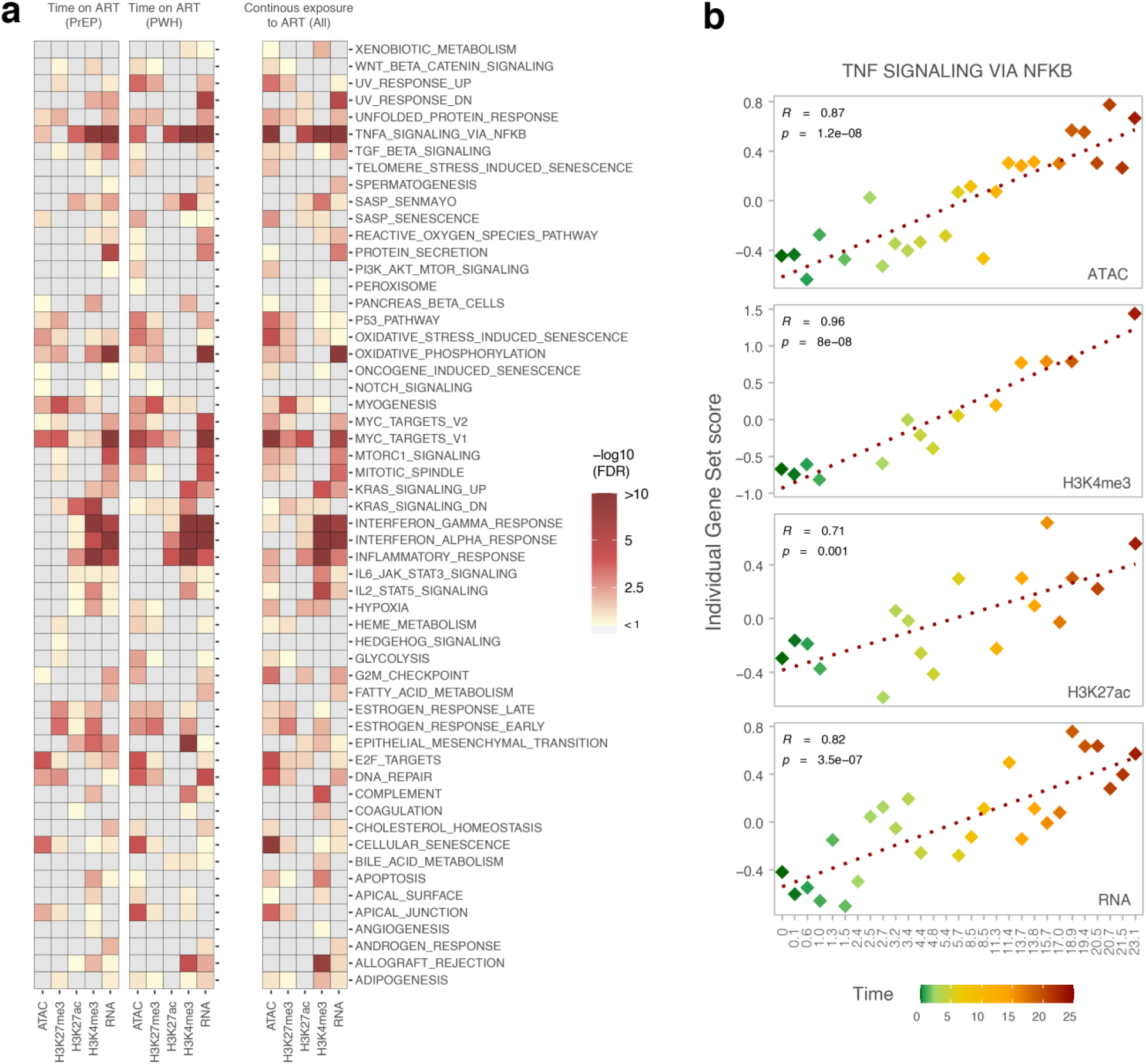
GESECA association with continuous exposure to. (**a**) GESECA for Hallmark pathways for time on ART in PrEP (left), in PWH (center), and continuous exposure to ART irrespective of the group (right). FDR for all tested gene sets is shown as a heatmap with increased significance from yellow to red.(**b**) Correlation of individual gene set scores (y-axis) for the TNF signaling pathway with length of ART exposure (x-axis). Pearson correlation R and corresponding *p-*values are shown at the top left of the plots.

**Fig. S7.**
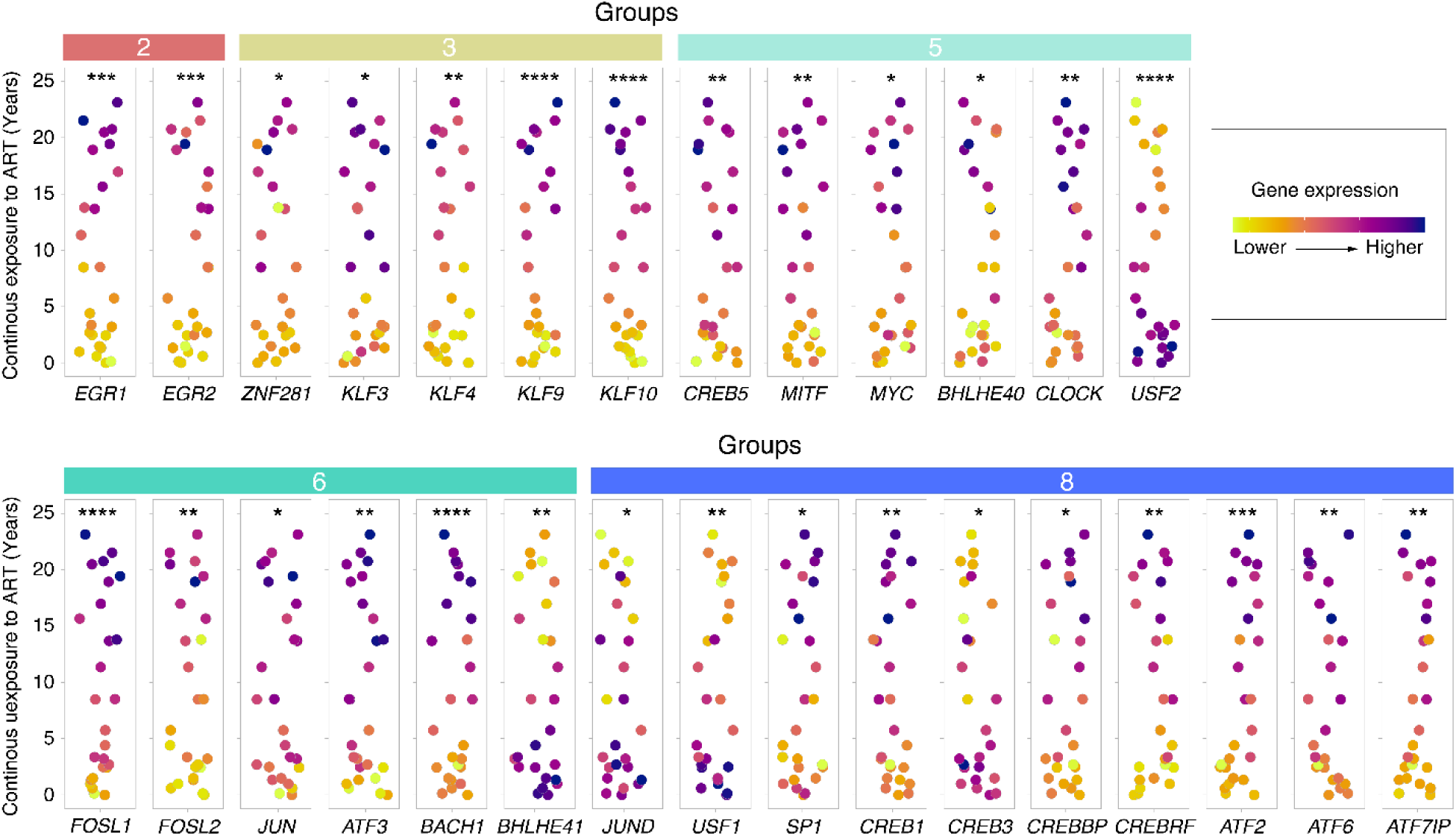
Coordinated increased gene expression in AM of transcription factors with motifs associated with continuous exposure to ART. Boxplots showing the time of continuous exposure to ART on the y-axes for genes encoding transcription factors (TF) shown in Fig. 4d. Each dot indicates the continuous exposure to ART for all subjects. The boxes at the top of the plots show the corresponding groups for the TF gene. FDR significance for the continuous exposure to ART. *, FDR < 10%, ** FDR < 1%, *** FDR < 0.1% and, **** FDR < 0.01%.

**Fig. S8.**
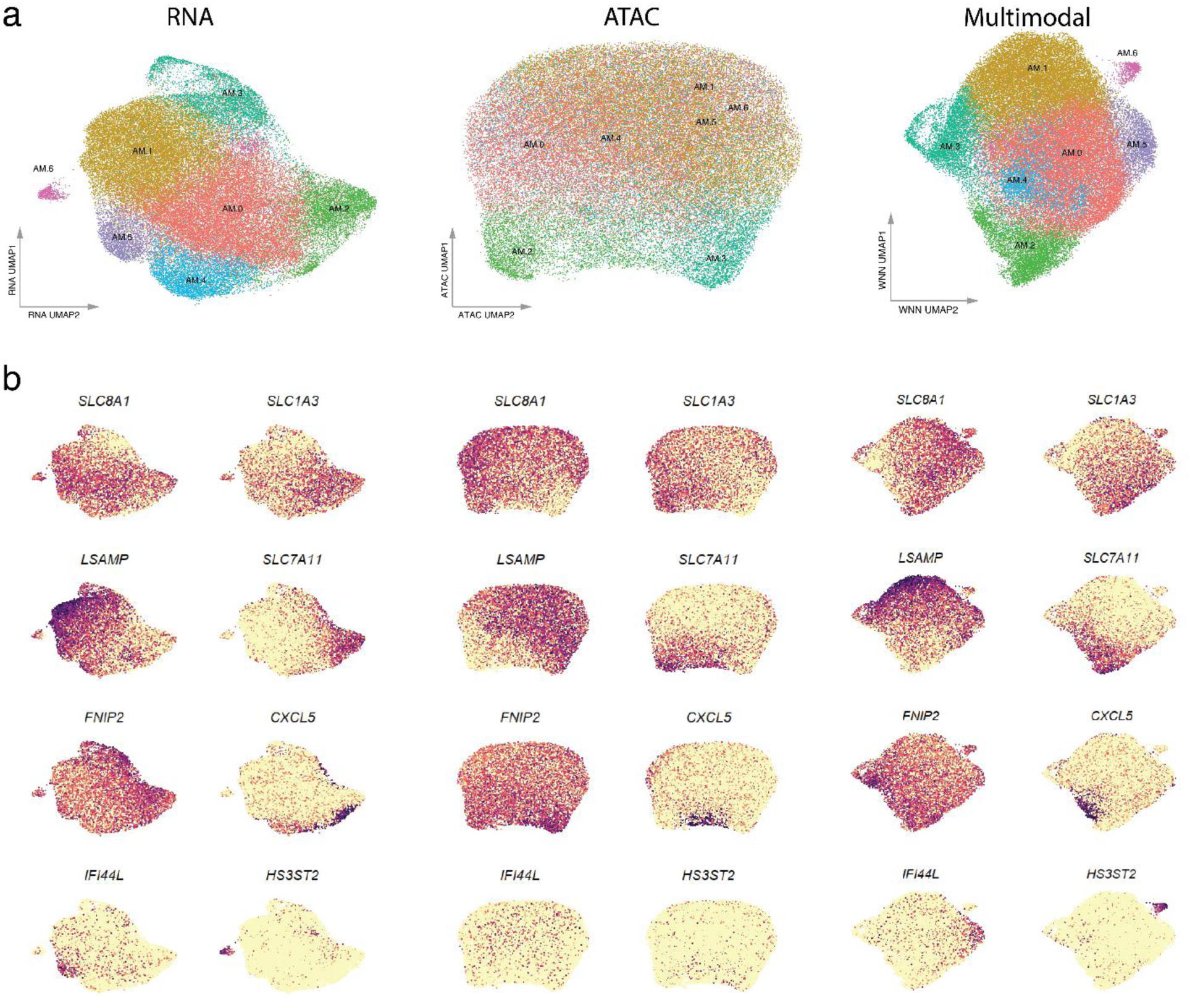
Annotation of AM subpopulation with single nucleus multiomics. (a) UMAPs for each modality (RNA and ATAC), as well as the multimodal weighted nearest neighbor (WNN) integration, showing the annotated AM subpopulations defined via single nucleus multimodality. (b) UMAPs displaying the expression of top cluster markers for each AM subpopulation.

**Fig S9.**
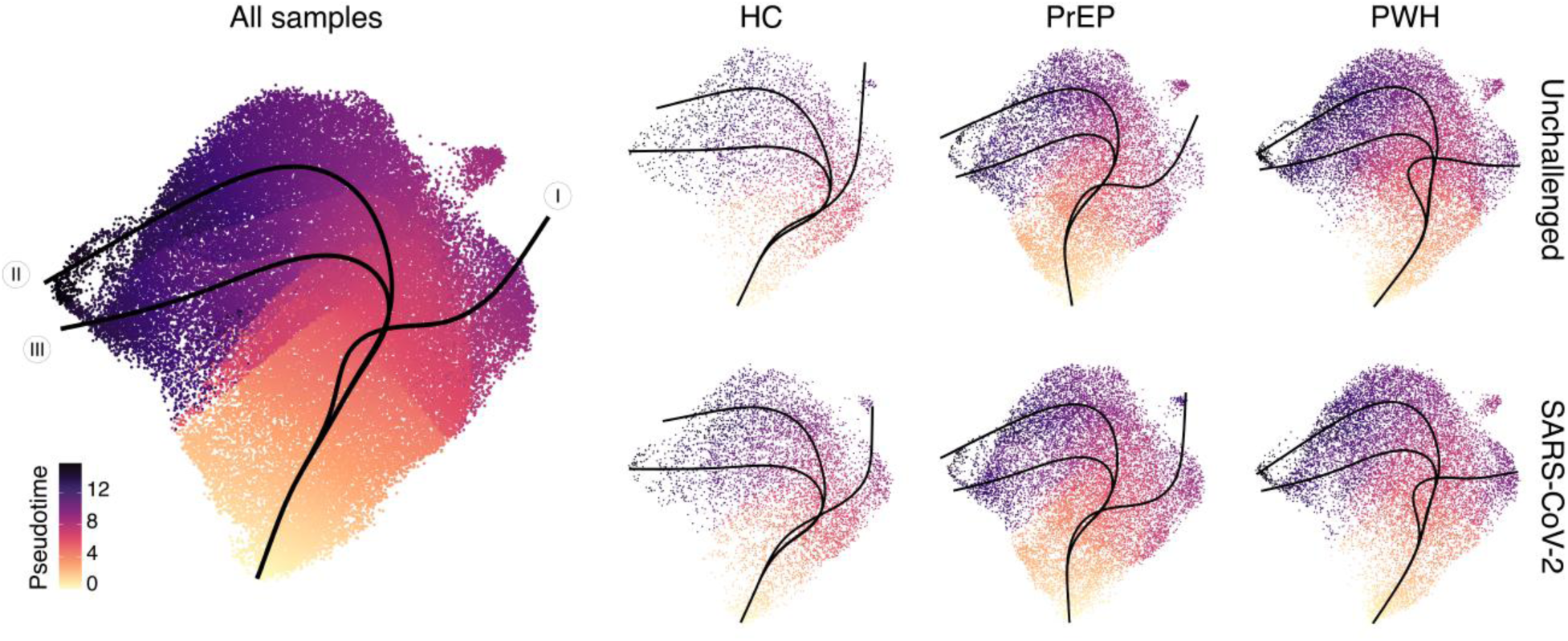
Pseudotime and trajectory analysis. The starting point of the pseudotime scale and the initial trajectory were obtained unsupervised from the data. Trajectory branches are shown as black lines on top of the UMAP. The left panel shows the trajectory obtained for all cells. The panel on the right shows the trajectory for each group separated by SARS-CoV-2 challenge status.

**Fig. S10.**
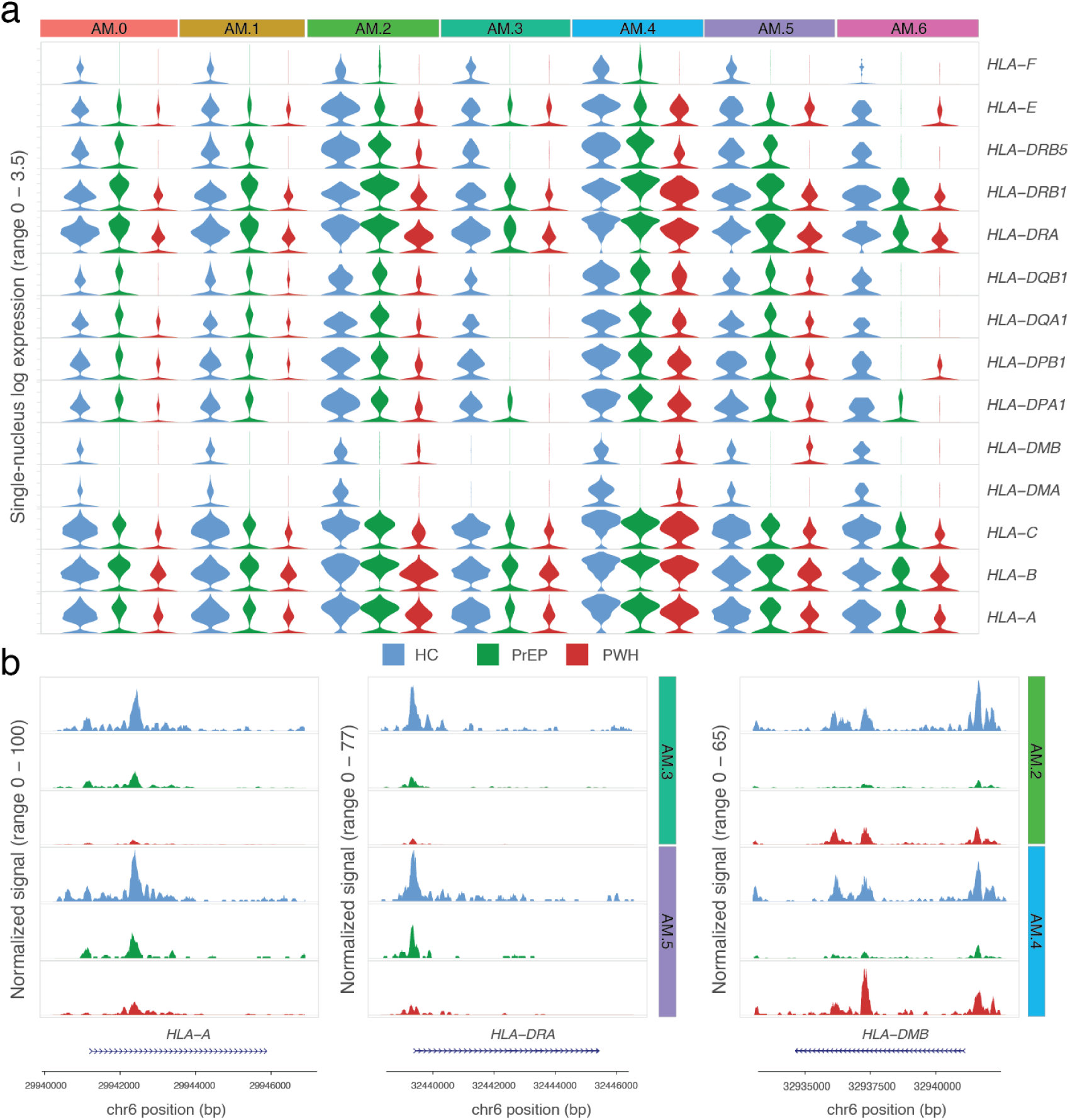
Constitutive HLA gene expression and chromatin accessibility is impaired in AM from participants on ART. (a) Single-nucleus expression of HLA genes. Violin plots indicate the per cell expression of the genes listed on the right. (b) Chromatin accessibility plots for the classical class I *HLA-A* and class II *HLA-DRB1* genes in the AM.3 and AM.5 subpopulations and the non-classical class II *HLA-DMB* for AM.2 and AM.4 subpopulations. Tracks display peaks representing baseline chromatin accessibility down sampled to show the same number of cells per group.

**Fig. S11.**
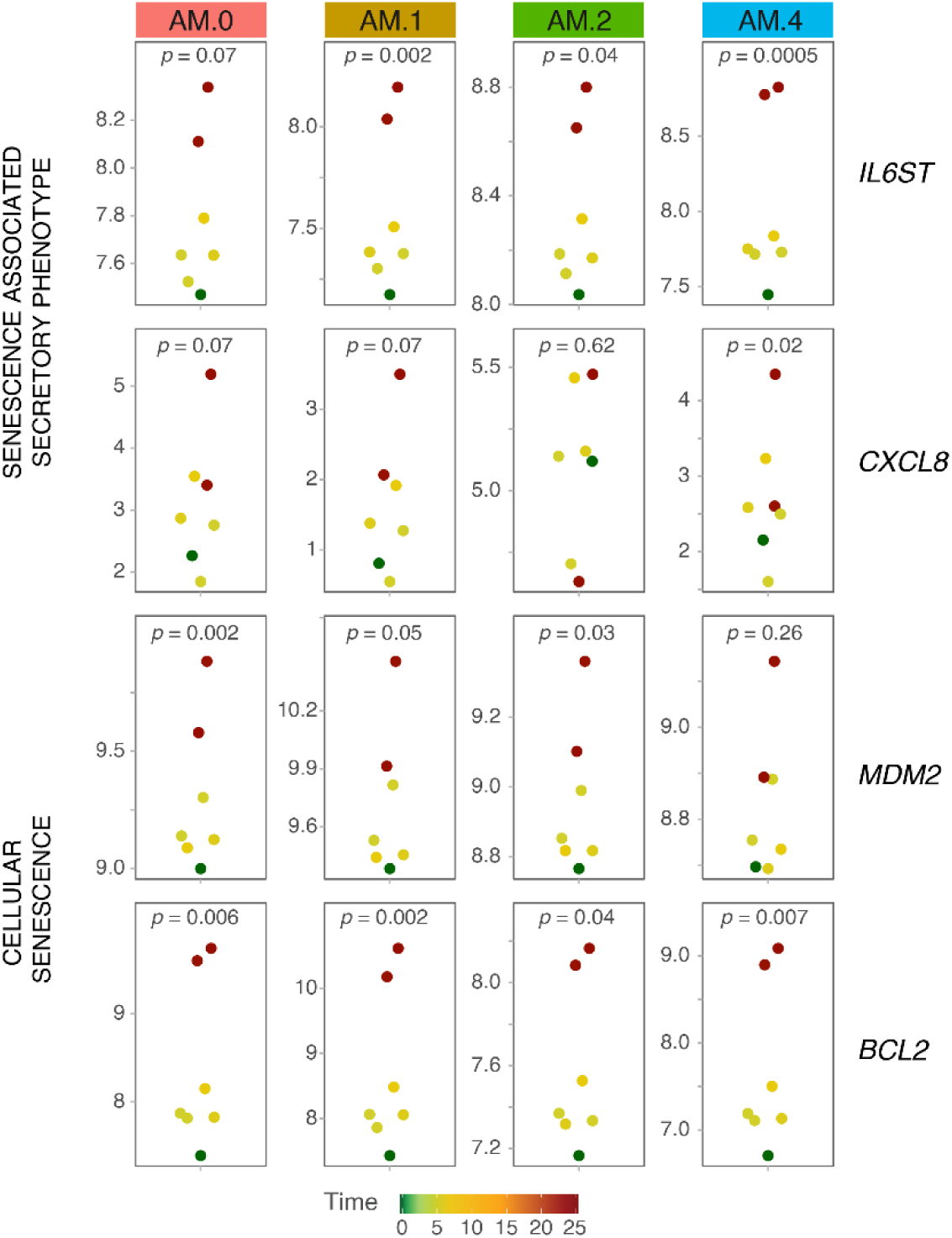
Senescence markers associate with continuous exposure to ART at the single-nucleus level. Normalized pseudobulk gene expression across timepoints of continuous exposure to ART per AM subpopulation for selected genes from Senescence Associated Secretory Phenotype (SASP) and cellular senescence HALLMARK gene sets.

## Reference

1. Tseng A, Seet J, Phillips EJ. The evolution of three decades of antiretroviral therapy: challenges, triumphs and the promise of the future. Br J Clin Pharmacol. 2015;79(2):182–94.

2. Premeaux TA, Ndhlovu LC. Decrypting biological hallmarks of aging in people with HIV. Curr Opin HIV AIDS. 2023;18(5):237–45.

3. Balderson BH, Grothaus L, Harrison RG, McCoy K, Mahoney C, Catz S. Chronic illness burden and quality of life in an aging HIV population. AIDS Care. 2013;25(4):451–8.

4. Nanditha NGA, Paiero A, Tafessu HM, St-Jean M, McLinden T, Justice AC, et al. Excess burden of age-associated comorbidities among people living with HIV in British Columbia, Canada: a population-based cohort study. BMJ open. 2021;11(1):e041734.

5. Chawla A, Wang C, Patton C, Murray M, Punekar Y, de Ruiter A, et al. A Review of Long-Term Toxicity of Antiretroviral Treatment Regimens and Implications for an Aging Population. Infect Dis Ther. 2018;7(2):183–95.

6. Fitzpatrick ME, Kunisaki KM, Morris A. Pulmonary disease in HIV-infected adults in the era of antiretroviral therapy. AIDS. 2018;32(3):277–92.

7. Crothers K, Huang L, Goulet JL, Goetz MB, Brown ST, Rodriguez-Barradas MC, et al. HIV infection and risk for incident pulmonary diseases in the combination antiretroviral therapy era. Am J Respir Crit Care Med. 2011;183(3):388–95.

8. Schoepf IC, Esteban-Cantos A, Thorball CW, Rodes B, Reiss P, Rodriguez-Centeno J, et al. Epigenetic ageing accelerates before antiretroviral therapy and decelerates after viral suppression in people with HIV in Switzerland: a longitudinal study over 17 years. Lancet Healthy Longev. 2023;4(5):e211–e8.

9. Dube M, Tastet O, Dufour C, Sannier G, Brassard N, Delgado GG, et al. Spontaneous HIV expression during suppressive ART is associated with the magnitude and function of HIV-specific CD4(+) and CD8(+) T cells. Cell Host Microbe. 2023;31(9):1507–22 e5.

10. Fastenackels S, Sauce D, Vigouroux C, Avettand-Fenoel V, Bastard JP, Fellahi S, et al. HIV-mediated immune aging in young adults infected perinatally or during childhood. AIDS. 2019;33(11):1705–10.

11. Yang CX, Schon E, Obeidat M, Kobor MS, McEwen L, MacIsaac J, et al. Occurrence of Accelerated Epigenetic Aging and Methylation Disruptions in Human Immunodeficiency Virus Infection Before Antiretroviral Therapy. J Infect Dis. 2021;223(10):1681–9.

12. Corley MJ, Sacdalan C, Pang APS, Chomchey N, Ratnaratorn N, Valcour V, et al. Abrupt and altered cell-type specific DNA methylation profiles in blood during acute HIV infection persists despite prompt initiation of ART. PLoS Pathog. 2021;17(8):e1009785.

13. Bigna JJ, Kenne AM, Asangbeh SL, Sibetcheu AT. Prevalence of chronic obstructive pulmonary disease in the global population with HIV: a systematic review and meta-analysis. Lancet Glob Health. 2018;6(2):e193–e202.

14. Wang Y, Xie Y, Hu S, Ai W, Tao Y, Tang H, et al. Systematic Review and Meta-Analyses of The Interaction Between HIV Infection And COVID-19: Two Years’ Evidence Summary. Front Immunol. 2022;13:864838.

15. Bertagnolio S, Thwin SS, Silva R, Nagarajan S, Jassat W, Fowler R, et al. Clinical features of, and risk factors for, severe or fatal COVID-19 among people living with HIV admitted to hospital: analysis of data from the WHO Global Clinical Platform of COVID-19. Lancet HIV. 2022;9(7):e486–e95.

16. WHO. Global tuberculosis report 2023. 2023.

17. Correa-Macedo W, Fava VM, Orlova M, Cassart P, Olivenstein R, Sanz J, et al. Alveolar macrophages from persons living with HIV show impaired epigenetic response to Mycobacterium tuberculosis. J Clin Invest. 2021;131(22).

18. Feikin DR, Feldman C, Schuchat A, Janoff EN. Global strategies to prevent bacterial pneumonia in adults with HIV disease. Lancet Infect Dis. 2004;4(7):445–55.

19. Zifodya JS, Crothers K. Treating bacterial pneumonia in people living with HIV. Expert Rev Respir Med. 2019;13(8):771–86.

20. Maitre T, Cottenet J, Beltramo G, Georges M, Blot M, Piroth L, et al. Increasing burden of noninfectious lung disease in persons living with HIV: a 7-year study using the French nationwide hospital administrative database. Eur Respir J. 2018;52(3).

21. Haas CB, Engels EA, Horner MJ, Freedman ND, Luo Q, Gershman S, et al. Trends and risk of lung cancer among people living with HIV in the USA: a population-based registry linkage study. Lancet HIV. 2022;9(10):e700–e8.

22. Sigel K, Makinson A, Thaler J. Lung cancer in persons with HIV. Curr Opin HIV AIDS. 2017;12(1):31–8.

23. Meyer KC, Raghu G, Baughman RP, Brown KK, Costabel U, du Bois RM, et al. An official American Thoracic Society clinical practice guideline: the clinical utility of bronchoalveolar lavage cellular analysis in interstitial lung disease. Am J Respir Crit Care Med. 2012;185(9):1004–14.

24. Parks B, Greenleaf W. Scalable high-performance single cell data analysis with BPCells. bioRxiv. 2025:2025.03.27.645853.

25. Hao Y, Stuart T, Kowalski MH, Choudhary S, Hoffman P, Hartman A, et al. Dictionary learning for integrative, multimodal and scalable single-cell analysis. Nature biotechnology. 2024;42(2):293–304.

26. Stuart T, Srivastava A, Madad S, Lareau CA, Satija R. Single-cell chromatin state analysis with Signac. Nature methods. 2021;18(11):1333–41.

27. Yu G, Wang LG, Han Y, He QY. clusterProfiler: an R package for comparing biological themes among gene clusters. OMICS. 2012;16(5):284–7.

28. Alquicira-Hernandez J, Powell JE. Nebulosa recovers single-cell gene expression signals by kernel density estimation. Bioinformatics. 2021;37(16):2485–7.

29. Street K, Risso D, Fletcher RB, Das D, Ngai J, Yosef N, et al. Slingshot: cell lineage and pseudotime inference for single-cell transcriptomics. BMC genomics. 2018;19(1):477.

30. Murall CL, Fournier E, Galvez JH, N’Guessan A, Reiling SJ, Quirion PO, et al. A small number of early introductions seeded widespread transmission of SARS-CoV-2 in Quebec, Canada. Genome Med. 2021;13(1):169.

31. Ritchie ME, Phipson B, Wu D, Hu Y, Law CW, Shi W, et al. limma powers differential expression analyses for RNA-sequencing and microarray studies. Nucleic Acids Res. 2015;43(7):e47.

32. Law CW, Chen Y, Shi W, Smyth GK. voom: Precision weights unlock linear model analysis tools for RNA-seq read counts. Genome Biol. 2014;15(2):R29.

33. Robinson MD, McCarthy DJ, Smyth GK. edgeR: a Bioconductor package for differential expression analysis of digital gene expression data. Bioinformatics. 2010;26(1):139–40.

34. Yu G, Wang LG, He QY. ChIPseeker: an R/Bioconductor package for ChIP peak annotation, comparison and visualization. Bioinformatics. 2015;31(14):2382–3.

35. Sergushichev AA. An algorithm for fast preranked gene set enrichment analysis using cumulative statistic calculation. bioRxiv. 2016:060012.

36. Liberzon A, Birger C, Thorvaldsdottir H, Ghandi M, Mesirov JP, Tamayo P. The Molecular Signatures Database (MSigDB) hallmark gene set collection. Cell Syst. 2015;1(6):417–25.

37. Milacic M, Beavers D, Conley P, Gong C, Gillespie M, Griss J, et al. The Reactome Pathway Knowledgebase 2024. Nucleic Acids Res. 2024;52(D1):D672–D8.

38. Saul D, Kosinsky RL, Atkinson EJ, Doolittle ML, Zhang X, LeBrasseur NK, et al. A new gene set identifies senescent cells and predicts senescence-associated pathways across tissues. Nature communications. 2022;13(1):4827.

39. Heinz S, Benner C, Spann N, Bertolino E, Lin YC, Laslo P, et al. Simple combinations of lineage-determining transcription factors prime cis-regulatory elements required for macrophage and B cell identities. Molecular cell. 2010;38(4):576–89.

40. Mahony S, Auron PE, Benos PV. DNA familial binding profiles made easy: comparison of various motif alignment and clustering strategies. PLoS Comput Biol. 2007;3(3):e61.

41. Bentsen M, Goymann P, Schultheis H, Klee K, Petrova A, Wiegandt R, et al. ATAC-seq footprinting unravels kinetics of transcription factor binding during zygotic genome activation. Nature communications. 2020;11(1):4267.

42. Zou J, Ernst J. Chromatin state modeling across individuals reveals global patterns of histone modifications. bioRxiv. 2022:2022.08.02.502571.

43. Grubert F, Zaugg JB, Kasowski M, Ursu O, Spacek DV, Martin AR, et al. Genetic Control of Chromatin States in Humans Involves Local and Distal Chromosomal Interactions. Cell. 2015;162(5):1051–65.

44. Ramirez F, Ryan DP, Gruning B, Bhardwaj V, Kilpert F, Richter AS, et al. deepTools2: a next generation web server for deep-sequencing data analysis. Nucleic Acids Res. 2016;44(W1):W160–5.

45. Rauluseviciute I, Riudavets-Puig R, Blanc-Mathieu R, Castro-Mondragon JA, Ferenc K, Kumar V, et al. JASPAR 2024: 20th anniversary of the open-access database of transcription factor binding profiles. Nucleic Acids Res. 2024;52(D1):D174-D82.

46. Wickham H. ggplot2: Elegant Graphics for Data Analysis: Springer Publishing Company, Incorporated; 2009. 216 p.

47. Kaplan D, Pruim R. ggformula: Formula Interface to the Grammar of Graphics. https://githubcom/ProjectMOSAIC/ggformula. 2023.

48. Badia IMP, Velez Santiago J, Braunger J, Geiss C, Dimitrov D, Muller-Dott S, et al. decoupleR: ensemble of computational methods to infer biological activities from omics data. Bioinform Adv. 2022;2(1):vbac016.

49. Dallmann-Sauer M, Fava VM, Malherbe ST, MacDonald CE, Orlova M, Kroon EE, et al. Mycobacterium tuberculosis resisters despite HIV exhibit activated T cells and macrophages in their pulmonary alveoli. J Clin Invest. 2025;135(7).

50. Fleck JS, Jansen SMJ, Wollny D, Zenk F, Seimiya M, Jain A, et al. Inferring and perturbing cell fate regulomes in human brain organoids. Nature. 2023;621(7978):365-72.

51. Fornes O, Castro-Mondragon JA, Khan A, van der Lee R, Zhang X, Richmond PA, et al. JASPAR 2020: update of the open-access database of transcription factor binding profiles. Nucleic Acids Res. 2020;48(D1):D87–D92.

52. Charrier-Savournin FB, Chateau MT, Gire V, Sedivy J, Piette J, Dulic V. p21-Mediated nuclear retention of cyclin B1-Cdk1 in response to genotoxic stress. Mol Biol Cell. 2004;15(9):3965–76.

53. Wang XW, Zhan Q, Coursen JD, Khan MA, Kontny HU, Yu L, et al. GADD45 induction of a G2/M cell cycle checkpoint. Proc Natl Acad Sci U S A. 1999;96(7):3706–11.

54. Zannini L, Delia D, Buscemi G. CHK2 kinase in the DNA damage response and beyond. J Mol Cell Biol. 2014;6(6):442–57.

55. Acosta JC, Banito A, Wuestefeld T, Georgilis A, Janich P, Morton JP, et al. A complex secretory program orchestrated by the inflammasome controls paracrine senescence. Nat Cell Biol. 2013;15(8):978–90.

56. Vadaq N, van de Wijer L, van Eekeren LE, Koenen H, de Mast Q, Joosten LAB, et al. Targeted plasma proteomics reveals upregulation of distinct inflammatory pathways in people living with HIV. iScience. 2022;25(10):105089.

57. Ellwanger JH, Valverde-Villegas JM, Kaminski VL, de Medeiros RM, Almeida SEM, Santos BR, et al. Increased IL-8 levels in HIV-infected individuals who initiated ART with CD4(+) T cell counts <350 cells/mm(3) -A potential hallmark of chronic inflammation. Microbes Infect. 2020;22(9):474–80.

58. Cohen J, D’Agostino L, Wilson J, Tuzer F, Torres C. Astrocyte Senescence and Metabolic Changes in Response to HIV Antiretroviral Therapy Drugs. Front Aging Neurosci. 2017;9:281.

59. Konstantinidis I, Crothers K, Kunisaki KM, Drummond MB, Benfield T, Zar HJ, et al. HIV-associated lung disease. Nat Rev Dis Primers. 2023;9(1):39.

60. Coghill AE, Han X, Suneja G, Lin CC, Jemal A, Shiels MS. Advanced stage at diagnosis and elevated mortality among US patients with cancer infected with HIV in the National Cancer Data Base. Cancer. 2019;125(16):2868–76.

61. Ucero AC, Bakiri L, Roediger B, Suzuki M, Jimenez M, Mandal P, et al. Fra-2-expressing macrophages promote lung fibrosis in mice. J Clin Invest. 2019;129(8):3293–309.

62. Birnhuber A, Biasin V, Schnoegl D, Marsh LM, Kwapiszewska G. Transcription factor Fra-2 and its emerging role in matrix deposition, proliferation and inflammation in chronic lung diseases. Cell Signal. 2019;64:109408.

63. Rincon M, Irvin CG. Role of IL-6 in asthma and other inflammatory pulmonary diseases. Int J Biol Sci. 2012;8(9):1281–90.

64. Liu F, Zhang X, Du W, Du J, Chi Y, Sun B, et al. Diagnosis values of IL-6 and IL-8 levels in serum and bronchoalveolar lavage fluid for invasive pulmonary aspergillosis in chronic obstructive pulmonary disease. J Investig Med. 2021;69(7):1344–9.

65. Chang YP, Tsai YH, Chen YM, Huang KT, Lee CP, Hsu PY, et al. Upregulated microRNA-125b-5p in patients with asthma-COPD overlap mediates oxidative stress and late apoptosis via targeting IL6R/TRIAP1 signaling. Respir Res. 2024;25(1):64.

66. Lopez-Otin C, Blasco MA, Partridge L, Serrano M, Kroemer G. Hallmarks of aging: An expanding universe. Cell. 2023;186(2):243–78.

67. Barnes PJ. Oxidative stress-based therapeutics in COPD. Redox Biol. 2020;33:101544.

68. Casanova-Acebes M, Dalla E, Leader AM, LeBerichel J, Nikolic J, Morales BM, et al. Tissue-resident macrophages provide a pro-tumorigenic niche to early NSCLC cells. Nature. 2021;595(7868):578-84.

69. Dayanc B, Eris S, Gulfirat NE, Ozden-Yilmaz G, Cakiroglu E, Coskun Deniz OS, et al. Integrative multi-omics identifies AP-1 transcription factor as a targetable mediator of acquired osimertinib resistance in non-small cell lung cancer. Cell Death Dis. 2025;16(1):414.

70. Hou H, Sun D, Zhang X. The role of MDM2 amplification and overexpression in therapeutic resistance of malignant tumors. Cancer Cell Int. 2019;19:216.

71. Zong D, Gu J, Cavalcante GC, Yao W, Zhang G, Wang S, et al. BRD4 Levels Determine the Response of Human Lung Cancer Cells to BET Degraders That Potently Induce Apoptosis through Suppression of Mcl-1. Cancer Res. 2020;80(11):2380–93.

72. Alexandrova Y, Costiniuk CT, Jenabian MA. Pulmonary Immune Dysregulation and Viral Persistence During HIV Infection. Front Immunol. 2021;12:808722.

73. Wang Z, Li S, Huang B. Alveolar macrophages: Achilles’ heel of SARS-CoV-2 infection. Signal Transduct Target Ther. 2022;7(1):242.

74. Bain CC, Lucas CD, Rossi AG. Pulmonary macrophages and SARS-Cov2 infection. Int Rev Cell Mol Biol. 2022;367:1–28.

75. Magnen M, You R, Rao AA, Davis RT, Rodriguez L, Bernard O, et al. Immediate myeloid depot for SARS-CoV-2 in the human lung. Sci Adv. 2024;10(31):eadm8836.

76. Kotov DI, Lee OV, Fattinger SA, Langner CA, Guillen JV, Peters JM, et al. Early cellular mechanisms of type I interferon-driven susceptibility to tuberculosis. Cell. 2023;186(25):5536–53 e22.

77. Bastard P, Rosen LB, Zhang Q, Michailidis E, Hoffmann HH, Zhang Y, et al. Autoantibodies against type I IFNs in patients with life-threatening COVID-19. Science. 2020;370(6515).

78. Philippot Q, Fekkar A, Gervais A, Le Voyer T, Boers LS, Conil C, et al. Autoantibodies Neutralizing Type I IFNs in the Bronchoalveolar Lavage of at Least 10% of Patients During Life-Threatening COVID-19 Pneumonia. J Clin Immunol. 2023;43(6):1093–103.

79. Pilkington V, Hill A, Hughes S, Nwokolo N, Pozniak A. How safe is TDF/FTC as PrEP? A systematic review and meta-analysis of the risk of adverse events in 13 randomised trials of PrEP. J Virus Erad. 2018;4(4):215–24.

80. Cohen MS, Chen YQ, McCauley M, Gamble T, Hosseinipour MC, Kumarasamy N, et al. Antiretroviral Therapy for the Prevention of HIV-1 Transmission. N Engl J Med. 2016;375(9):830–9.

81. Marrazzo J, Tao L, Becker M, Leech AA, Taylor AW, Ussery F, et al. HIV Preexposure Prophylaxis With Emtricitabine and Tenofovir Disoproxil Fumarate Among Cisgender Women. JAMA. 2024;331(11):930–7.

82. Cox SN, Wu L, Wittenauer R, Clark S, Roberts DA, Nwogu IB, et al. Impact of HIV self-testing for oral pre-exposure prophylaxis scale-up on drug resistance and HIV outcomes in western Kenya: a modelling study. Lancet HIV. 2024;11(3):e167–e75.

83. Choopanya K, Martin M, Suntharasamai P, Sangkum U, Mock PA, Leethochawalit M, et al. Antiretroviral prophylaxis for HIV infection in injecting drug users in Bangkok, Thailand (the Bangkok Tenofovir Study): a randomised, double-blind, placebo-controlled phase 3 trial. Lancet. 2013;381(9883):2083–90.

84. Baeten JM, Donnell D, Ndase P, Mugo NR, Campbell JD, Wangisi J, et al. Antiretroviral prophylaxis for HIV prevention in heterosexual men and women. N Engl J Med. 2012;367(5):399–410.

85. Jourdain H, de Gage SB, Desplas D, Dray-Spira R. Real-world effectiveness of pre-exposure prophylaxis in men at high risk of HIV infection in France: a nested case-control study. Lancet Public Health. 2022;7(6):e529–e36.

86. Hughes SM, Levy CN, Calienes FL, Stekler JD, Pandey U, Vojtech L, et al. Treatment with Commonly Used Antiretroviral Drugs Induces a Type I/III Interferon Signature in the Gut in the Absence of HIV Infection. Cell Rep Med. 2020;1(6):100096.

87. Bowman ER, Cameron C, Richardson B, Kulkarni M, Gabriel J, Kettelhut A, et al. In Vitro Exposure of Leukocytes to HIV Preexposure Prophylaxis Decreases Mitochondrial Function and Alters Gene Expression Profiles. Antimicrob Agents Chemother. 2020;65(1).

88. Cohen J, D’Agostino L, Tuzer F, Torres C. HIV antiretroviral therapy drugs induce premature senescence and altered physiology in HUVECs. Mech Ageing Dev. 2018;175:74–82.

89. Wallace J, Gonzalez H, Rajan R, Narasipura SD, Virdi AK, Olali AZ, et al. Anti-HIV Drugs Cause Mitochondrial Dysfunction in Monocyte-Derived Macrophages. Antimicrob Agents Chemother. 2022;66(4):e0194121.

90. Leeansyah E, Cameron PU, Solomon A, Tennakoon S, Velayudham P, Gouillou M, et al. Inhibition of telomerase activity by human immunodeficiency virus (HIV) nucleos(t)ide reverse transcriptase inhibitors: a potential factor contributing to HIV-associated accelerated aging. J Infect Dis. 2013;207(7):1157–65.

91. Kim S, Lee SA, Yoon H, Kim MY, Yoo JK, Ahn SH, et al. Exosome-based delivery of super-repressor IkappaBalpha ameliorates kidney ischemia-reperfusion injury. Kidney Int. 2021;100(3):570–84.

92. Park YJ, Bae J, Yoo JK, Ahn SH, Park SY, Kim YS, et al. Effects of NF-kappaB Inhibitor on Sepsis Depend on the Severity and Phase of the Animal Sepsis Model. J Pers Med. 2024;14(6).

93. Lee CJ, Jang SH, Lim J, Park H, Ahn SH, Park SY, et al. Exosome-based targeted delivery of NF-kappaB ameliorates age-related neuroinflammation in the aged mouse brain. Exp Mol Med. 2025;57(1):235–48.

94. Yuan X, Ruan W, Bobrow B, Carmeliet P, Eltzschig HK. Targeting hypoxia-inducible factors: therapeutic opportunities and challenges. Nat Rev Drug Discov. 2024;23(3):175–200.

95. Bui BP, Nguyen PL, Lee K, Cho J. Hypoxia-Inducible Factor-1: A Novel Therapeutic Target for the Management of Cancer, Drug Resistance, and Cancer-Related Pain. Cancers (Basel). 2022;14(24).

96. Wicks EE, Semenza GL. Hypoxia-inducible factors: cancer progression and clinical translation. J Clin Invest. 2022;132(11).

97. Chaib S, Tchkonia T, Kirkland JL. Cellular senescence and senolytics: the path to the clinic. Nat Med. 2022;28(8):1556–68.

98. Mannarino M, Cherif H, Ghazizadeh S, Martinez OW, Sheng K, Cousineau E, et al. Senolytic treatment for low back pain. Sci Adv. 2025;11(11):eadr1719.

99. Rivera CG, Zeuli JD, Smith BL, Johnson TM, Bhatia R, Otto AO, et al. HIV Pre-Exposure Prophylaxis: New and Upcoming Drugs to Address the HIV Epidemic. Drugs. 2023;83(18):1677–98.

100. Landovitz RJ, Donnell D, Clement ME, Hanscom B, Cottle L, Coelho L, et al. Cabotegravir for HIV Prevention in Cisgender Men and Transgender Women. N Engl J Med. 2021;385(7):595–608.

101. Delany-Moretlwe S, Hughes JP, Bock P, Ouma SG, Hunidzarira P, Kalonji D, et al. Cabotegravir for the prevention of HIV-1 in women: results from HPTN 084, a phase 3, randomised clinical trial. Lancet. 2022;399(10337):1779–89.

102. Marrazzo J. Lenacapavir for HIV-1 -Potential Promise of a Long-Acting Antiretroviral Drug. N Engl J Med. 2022;386(19):1848–9.

